# Clinical Effects of Low-Dose Esketamine During Emergency Conversion From Labor Epidural Analgesia to Category I Cesarean Section: A Randomized Controlled Trial of Maternal Hemodynamics, Analgesia, Sedation, and Neonatal Outcomes

**DOI:** 10.64898/2026.10.10.26365195

**Authors:** Weiyun Liang, Liping Zhai

## Abstract

**Background:** Emergency conversion from labor epidural analgesia to Category I cesarean section leaves little time for epidural top-up to produce a surgical block. We examined whether low-dose intravenous esketamine could provide short-term bridging analgesia and sedation while maintaining maternal hemodynamics, oxygenation, and early neonatal safety.

**Methods:** In this randomized study, parturients requiring Category I cesarean section after labor epidural analgesia received either esketamine 0.15 mg/kg or normal saline 1 min before skin incision, together with epidural alkalinized lidocaine. The primary outcome was the longitudinal change in maternal systolic blood pressure during the 20-min observation window. Secondary outcomes included heart rate, SpO_2_, intraoperative pain, surgeon satisfaction with muscle relaxation, Ramsay sedation score, and neonatal Apgar scores. We analyzed repeated continuous outcomes with linear mixed-effects models and repeated binary outcomes with generalized estimating equations. The single post-administration Ramsay outcome was analyzed using logistic regression. Apgar scores were summarized descriptively using medians, interquartile ranges, and counts.

**Results:** We analyzed 55 parturients, 31 in the esketamine group and 24 in the normal saline group. Esketamine did not meaningfully alter the systolic blood pressure trajectory. Heart rate and SpO_2_ showed broadly similar patterns between groups, with no signal of esketamine-related respiratory deterioration. The esketamine group had lower odds of intraoperative pain at 5, 15, and 20 min and a higher probability of target Ramsay sedation. Surgeon satisfaction with muscle relaxation showed no clear between-group difference. Apgar scores remained high in both groups.

**Conclusion:** Low-dose intravenous esketamine 0.15 mg/kg may be a practical bridging adjunct during emergency conversion from labor epidural analgesia to Category I cesarean section. It improved early pain control and target sedation without a clear adverse signal in systolic blood pressure, heart rate, oxygenation, or early Apgar scores. Trial Registration: Chinese Clinical Trial Registry, ChiCTR2600121948; retrospectively registered.

## Introduction

Vaginal delivery remains the predominant mode of childbirth, and neuraxial labor analgesia is widely used because of its reliable analgesic effect and favorable safety profile [1]. The epidural catheter left in place during labor analgesia may also provide an important anesthetic route for emergency conversion to cesarean section during labor. When obstetric emergencies threaten maternal or fetal safety, a Category I cesarean section must be performed without delay. In this setting, the decision-to-delivery interval (DDI) is generally expected to be within 30 min, and a shorter interval is associated with better maternal and fetal outcomes [2].

In clinical practice, however, emergency conversion from labor analgesia to cesarean section often creates a conflict between the onset time of epidural local anesthetics and the need for rapid fetal delivery. During labor analgesia, repeated epidural administration of low-concentration local anesthetics may contribute to delayed onset or incomplete block when high-concentration local anesthetics are subsequently administered [3]. Even with alkalinized local anesthetics, which are used to accelerate onset, an adequate surgical anesthetic level may still not be achieved quickly enough in highly urgent situations [4]. General anesthesia is also not an ideal rescue option in many parturients. Because oral intake during labor is often allowed to preserve maternal strength, many patients are in a full-stomach state, increasing the risk of aspiration during general anesthesia [5]. If inadequate neuraxial anesthesia is recognized only after surgery has begun, subsequent conversion to general anesthesia may further delay fetal delivery.

These circumstances create a narrow clinical window in which neuraxial block is not yet fully established, but fetal delivery cannot be postponed. A short-acting adjunct capable of providing analgesia and appropriate sedation before the epidural top-up has taken full effect may therefore be clinically useful. However, sedative or analgesic drugs given before fetal delivery must be used cautiously, because maternal hemodynamic instability and reduced uteroplacental perfusion may worsen outcomes, particularly in obstetric emergencies such as placental abruption or fetal distress [6]. Possible fetal exposure through placental transfer is another important consideration when an intravenous adjunct is administered before delivery.

Ketamine has potential advantages in this setting because it provides analgesia with relatively limited respiratory depression and can help maintain circulatory stability [7–8]. Esketamine, the dextrorotatory isomer of ketamine, has higher affinity for NMDA receptors and greater analgesic potency, allowing effective dosing at lower doses than racemic ketamine [9–10]. Although subanesthetic-dose ketamine has been used in various perioperative settings [8], evidence regarding esketamine in obstetric anesthesia remains limited, especially during emergency conversion from labor analgesia to Category I cesarean section [11–12].

On this basis, the present study evaluated low-dose intravenous esketamine combined with epidural alkalinized lidocaine in parturients undergoing emergency conversion from labor analgesia to Category I cesarean section. We focused primarily on maternal hemodynamic stability and also assessed analgesia, sedation, surgeon satisfaction with muscle relaxation, oxygenation, and early neonatal outcomes.

## Materials and Methods

### Study design and ethics

This single-center, randomized, double-blind, parallel-group controlled trial based on unrestricted simple randomization planned 1:1 allocation was conducted at Foshan Nanhai District Maternal and Child Health Hospital, Foshan, Guangdong, China. The authors confirm that all ongoing and related trials for this drug/intervention are registered.Interventions were delivered by anesthesiologists experienced in obstetric anesthesia. Participants were enrolled between 22 September 2023 and 1 May 2026. The first follow-up assessments were conducted between 23 September 2023 and 2 May 2026, and the second follow-up assessments were conducted between 2 October 2023 and 11 May 2026.The trial was designed to evaluate the clinical effects and safety profile of low-dose esketamine compared with normal saline in this emergency obstetric anesthesia setting. The study was reviewed and approved by the Ethics Committee of Foshan Nanhai District Maternal and Child Health Hospital before participant enrollment (approval No. 2022-01) and was conducted in accordance with the ethical principles of the Declaration of Helsinki. The study was also subject to continuing ethics supervision and annual follow-up review by the Ethics Committee during the study period. Written informed consent was obtained from all participants. Written informed consent was obtained from all participants. The study design and reporting followed the CONSORT guidelines [13].The trial was registered at the Chinese Clinical Trial Registry (ChiCTR2600121948; registration date: April 7, 2026). Registration was completed after recruitment had begun because the investigators became aware of the public trial registration requirement during the study process,although ethics approval was obtained before enrollment and institutional oversight continued throughout the study.

### Protocol modifications

During study implementation, no changes were made to the randomization procedure, intervention regimen, or eligibility criteria. To standardize outcome assessment and optimize data collection, the primary and secondary outcomes were explicitly distinguished. Systolic blood pressure was defined as the primary outcome, whereas heart rate, peripheral oxygen saturation (SpO_2_), intraoperative pain, surgeon satisfaction with muscle relaxation, Ramsay sedation score, and neonatal Apgar score were defined as secondary outcomes.

Systolic blood pressure replaced mean arterial pressure as the primary hemodynamic outcome because systolic blood pressure is routinely used in clinical monitoring and directly reflects perioperative circulatory changes in this surgical setting. In addition, maternal SpO_2_ and intraoperative pain were added as secondary outcomes to further strengthen the evidence chain regarding maternal respiratory safety and analgesic efficacy. SpO_2_ provides an objective assessment of oxygenation, whereas intraoperative pain reflects the clinical analgesic effect during the short interval before the epidural top-up has taken full effect.

These refinements were intended to improve the clarity and completeness of outcome assessment. They did not affect participant allocation, the randomized controlled design, the intervention regimen, or the inclusion and exclusion criteria.

### Participants

Participants were eligible if all of the following criteria were met: immediate Category I cesarean section was required because of an obstetric emergency; pain was still present on preoperative testing of the intended skin incision area; gestational age was ≥37 weeks; estimated fetal weight was ≥2.5 kg; an epidural catheter had been successfully placed at the L2–3 interspace with an uneventful puncture procedure; the initial effect of labor analgesia was satisfactory (visual analogue scale [VAS] score 1–2); the last fetal heart rate before conversion to surgery was ≥120 beats/min; American Society of Anesthesiologists (ASA) physical status was I–II; and routine prenatal examinations showed no obvious abnormalities.

Participants were excluded if any of the following conditions were present: cardiovascular disease; recent use of analgesic drugs; psychiatric disorders preventing cooperation with the study procedures; hypertension; conversion to general anesthesia or repeat neuraxial puncture because of markedly inadequate anesthetic effect; conversion to vaginal delivery after entering the operating room; an interval of more than 5 min between the first epidural administration of high-concentration local anesthetic and skin incision; or the need for vasopressors or chronotropic agents because of a decrease in blood pressure or heart rate.

Because emergency conversion from labor epidural analgesia to Category I cesarean section is uncommon, highly time-sensitive, and difficult to predict, a formal efficacy-based sample size calculation was not performed before enrollment. At trial registration, the target enrollment was set at 70 parturients as a feasibility-based recruitment goal for the approved study period. With the planned 1:1 allocation, this target corresponded to approximately 35 participants per group and was expected to provide preliminary estimates of the variability in systolic blood pressure, the primary outcome, as well as other maternal and neonatal outcomes. Published guidance suggests that approximately 25 participants per group may be appropriate for an external pilot randomized trial when a small standardized effect is anticipated in a future definitive trial . All consecutive eligible parturients were screened during the study period, and 55 parturients completed the study procedures. The difference between the registered target and the final sample reflected the limited number of eligible cases in this emergency setting.

With the achieved sample, the pooled standard deviation of a continuous outcome could be estimated with an approximate 95% confidence interval ranging from 0.84 to 1.23 times the observed pooled standard deviation. Treatment-effect estimates for the longitudinal outcomes were therefore presented with 95% confidence intervals to show their precision. The study was intended to provide preliminary estimates and was not powered to draw definitive conclusions regarding efficacy or safety.[14]No interim analysis was planned or performed, and no formal stopping guidelines were prespecified.

### Randomization and blinding

Participants were randomly assigned to the normal saline group (NS group) or the esketamine group (KE group) using a computer-generated random allocation sequence based on unrestricted simple randomization with planned 1:1 allocation. The random allocation sequence was generated by an independent coordinator who was not involved in clinical anesthesia management, outcome assessment, or data analysis. Allocation concealment was maintained using sealed opaque envelopes, which were opened sequentially by the independent coordinator only after participant enrollment. Clinical personnel who enrolled participants had no access to the random allocation sequence. The independent coordinator was responsible for implementing the allocation sequence, opening the sealed opaque envelopes after enrollment, and preparing the assigned study syringe. According to group allocation, the independent coordinator prepared identical 10-mL syringes containing either esketamine or normal saline.The two study syringes were similar in appearance and volume, and the study drug was administered at 1min before operation . Patients, attending anesthesiologists who administered the study drug and provided intraoperative care, outcome assessors, and data analysts were blinded to group allocation throughout the study.

### Interventions

Before anesthesia, esketamine 50 mg was diluted with normal saline to a total volume of 10 mL. According to the randomization sequence, an independent coordinator prepared identical 10-mL syringes containing either esketamine or normal saline. After entry into the operating room, routine monitoring, including electrocardiography, heart rate, noninvasive blood pressure, and peripheral oxygen saturation, was established. The position of the epidural catheter at the back was checked, and catheter patency was confirmed before epidural drug administration.

All participants received epidural alkalinized lidocaine through the existing epidural catheter. The epidural solution consisted of 20 mL of 2% lidocaine mixed with 3 mL of 5% sodium bicarbonate. A 5-mL test dose was administered first. If no abnormal response occurred, an additional 10 – 15 mL was administered intermittently as needed until the sensory block reached the T6 dermatome.the operating table was tilted to the left to reduce the risk of supine hypotensive syndrome The study drug was administered intravenously 1 min before operation skin incision. Participants in the esketamine group received intravenous esketamine 0.15 mg/kg, whereas participants in the normal saline group received an equivalent weight-adjusted volume of intravenous normal saline. The attending anesthesiologist remained blinded to group allocation.Refer to Figure 1 for the detailed operational procedure.

**Figure 1.**
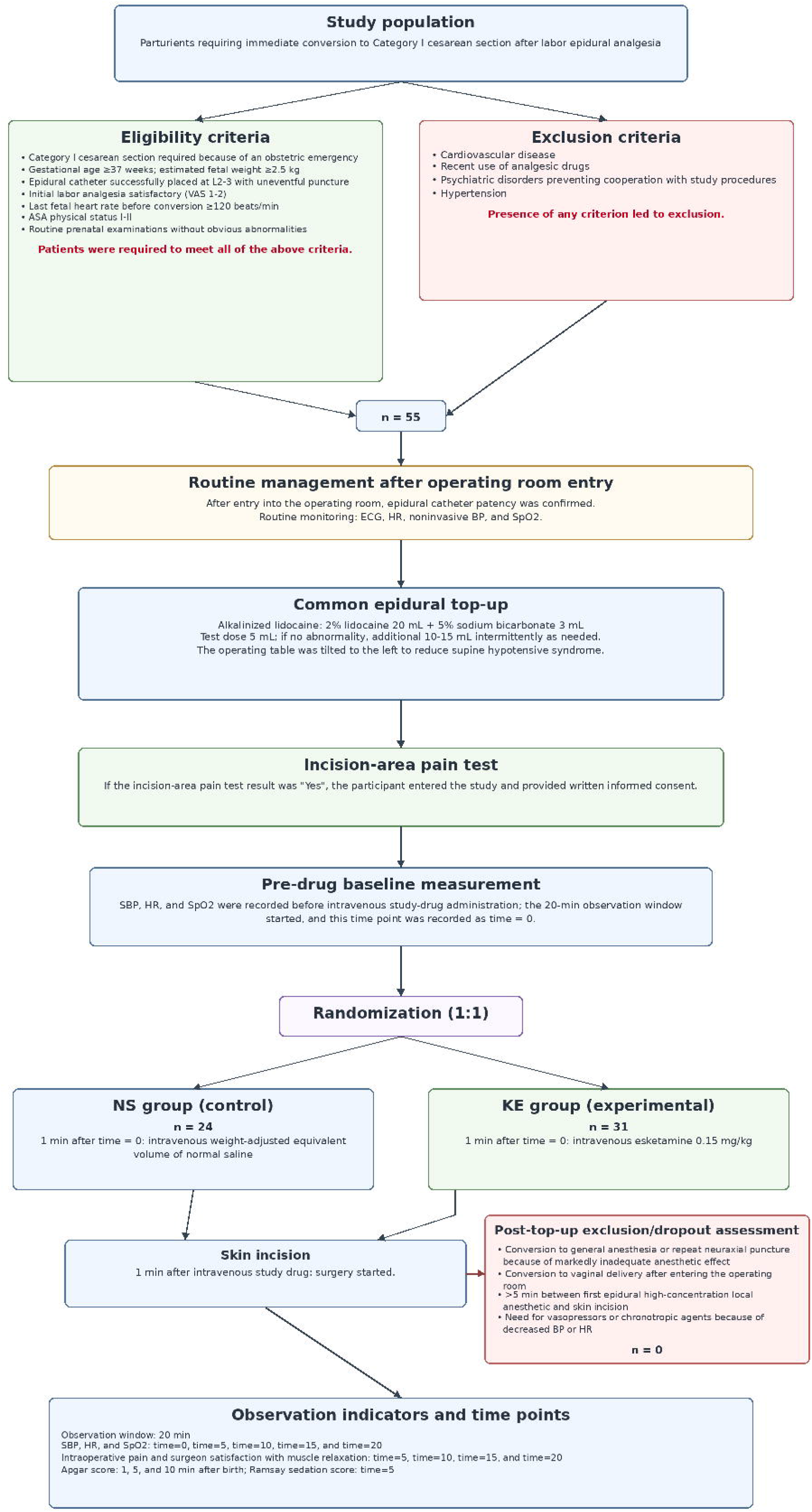
Study Flowchart

### Outcomes and measurements

Primary outcome:

Systolic Blood Pressure (changes in maternal systolic blood pressure): Maternal systolic blood pressure was recorded starting 1 min before intravenous administration and then measured every 5 min thereafter, for a total of 5 assessments.

These assessments corresponded to 0, 5, 10, 15, and 20 min during the observation window. Systolic blood pressure was recorded in mmHg and was analyzed as a repeated continuous outcome using longitudinal change over time.

Secondary outcomes:

Heart Rate (changes in maternal heart rate): Maternal heart rate was recorded starting 1 min before intravenous administration and then measured every 5 min thereafter, for a total of 5 assessments.

These assessments corresponded to 0, 5, 10, 15, and 20 min during the observation window. Heart rate was recorded in beats/min and was analyzed as a repeated continuous outcome using longitudinal change over time.

SpO_2_ (changes in maternal oxygen saturation): Maternal oxygen saturation (SpO_2_) was recorded starting 1 min before intravenous administration and then measured every 5 min thereafter, for a total of 5 assessments.

These assessments corresponded to 0, 5, 10, 15, and 20 min during the observation window. SpO_2_ was expressed as a percentage and was analyzed as a repeated continuous outcome using longitudinal change over time.

For the repeated continuous outcomes, the 0 min measurement were collected before drug administration and were therefore considered the baseline value for the corresponding variable, including baseline systolic blood pressure, baseline heart rate, and baseline SpO2.

Apgar Score [16]: Neonatal Apgar scores were recorded at 1, 5, and 10 min after birth, for a total of 3 assessments.

Because Apgar scores showed limited variability and a ceiling effect, they were summarized descriptively using medians, interquartile ranges, and counts.

Hurt (whether intraoperative pain was present): The presence or absence of maternal intraoperative pain was recorded every 5 min beginning at the start of surgery, for a total of 4 assessments.

These assessments corresponded to 5, 10, 15, and 20 min. Intraoperative pain was recorded as a binary outcome, with 0 indicating no pain and 1 indicating pain. This outcome was analyzed as a repeated binary outcome, and treatment effects were summarized using odds ratios with 95% confidence intervals. Absolute event rates were presented as n/N (%), where n represents the number of participants with intraoperative pain and N represents the number of participants with available data at the corresponding time point.

Anesthesia satisfaction (surgeon satisfaction with the degree of muscle relaxation): Whether the surgeon was satisfied with the degree of muscle relaxation was recorded every 5 min from the start of surgery, for a total of 4 assessments.

These assessments corresponded to 5, 10, 15, and 20 min. Surgeon satisfaction with muscle relaxation was recorded as a binary outcome, with 0 indicating not satisfied and 1 indicating satisfied. This outcome was analyzed as a repeated binary outcome, and treatment effects were summarized using odds ratios with 95% confidence intervals. Absolute event rates were presented as n/N (%), where n represents the number of participants with surgeon satisfaction and N represents the number of participants with available data at the corresponding time point.

Ramsay Score [17] (sedation score): Maternal sedation was assessed once during the skin incision period(time = 5).for a total of 1 assessment.

The Ramsay sedation scale was used:

1 = Awake; agitated or restless or both;

2 = Awake; cooperative, oriented, and tranquil;

3 = Awake but responds to commands only;

4 = Asleep; brisk response to light glabellar tap or loud auditory stimulus;

5 = Asleep; sluggish response to light glabellar tap or loud auditory stimulus;

6 = Asleep; no response to glabellar tap or loud auditory stimulus.

Maternal sedation was assessed once using the Ramsay sedation scale. Because postoperative communication between medical staff and patients after fetal delivery could influence sedation assessment, only the first post-administration Ramsay score at 5 min was used for analysis. Ramsay score was analyzed as a binary outcome, with scores of 2–4 defined as target sedation and other scores defined as non-target sedation. Treatment effects were summarized using odds ratios with 95% confidence intervals. Absolute event rates were presented as n/N (%), where n represents the number of participants with target sedation and N represents the number of participants with available Ramsay sedation data.

(see the Supporting Information S1 Table and Data dictionary)

### Harms assessment

Potential maternal and neonatal harms were prespecified in the study protocol and monitored during the perioperative observation period. Maternal harms of interest included allergic reactions, local anesthetic systemic toxicity, excessive sedation or respiratory depression, high neuraxial block, hemodynamic instability requiring rescue treatment, conversion to general anesthesia, or repeat neuraxial puncture. Neonatal harm was assessed primarily using Apgar scores at 1, 5, and 10 min after birth.

Maternal hemodynamic and respiratory safety was assessed systematically using prespecified measurements of systolic blood pressure, heart rate, SpO_2_, and Ramsay sedation score. According to the study protocol, participants who developed clinically significant hypotension or bradycardia requiring vasopressor or chronotropic treatment were to receive immediate rescue treatment and were withdrawn from the study analysis. Other adverse clinical events and rescue interventions, if present, were recorded from perioperative monitoring and clinical records. No separate structured questionnaire was used to assess neuropsychological symptoms such as dizziness, hallucinations, or dysphoria.

### Statistical Analysis

All data processing and statistical analyses were performed using R software version 4.5.3.

Repeated continuous outcomes, including systolic blood pressure (SBP), heart rate(HR), and oxygen saturation (SpO_2_), we analysis them using linear mixed-effects models (LMMs). Fixed effects included treatment group(drug), time, and the group-by-time interaction, while baseline values(time = 0 values) of each outcome were included as covariates. A random intercept for each participant was specified to account for within-subject correlation due to repeated measurements. Baseline values were centered before inclusion in the corresponding models. The model structure was as follows:

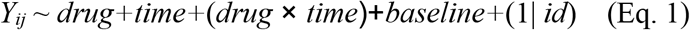

where*Y_ij_* represents the continuous outcome (SBP, HR, or SpO_2_) for participant *i* at time point *j*.

We using lmerTest package to fit the models, and *P*-values for fixed effects were calculated using Satterthwaite’s approximation.Because SpO_2_ values were bounded and has a ceiling effect, we fit it using beta mixed-effects model[18]as a sensitivity analysis after transformation to the open interval (0, 1).

To capture potential non-linear trajectories over time, we specified an expanded model incorporating quadratic time terms(*time^2^*) and their interaction with treatment group (Eq. 2):

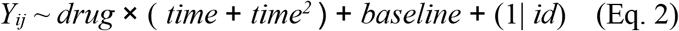

We systematically evaluated model fit between Eq. 1 and Eq. 2 using Likelihood Ratio Tests (LRT) under Maximum Likelihood (ML) estimation, alongside Akaike Information Criterion (AIC) and Bayesian Information Criterion (BIC), and estimated final parameters using Restricted Maximum Likelihood (REML). We verified Gaussian assumptions by inspecting Q-Q plots for conditional residuals and estimated random intercepts, and we prespecified Mixed-Effects Model with linear time term or Generalized Linear Mixed Models (GLMMs) as our sensitivity analysis if data substantially violated these normality assumptions.

Repeated binary outcomes, including Hurt and anesthesia satisfaction, we analysis them using generalized estimating equations (GEE) with a binomial distribution and logit link. And we use Robust sandwich variance estimators to calculate standard errors, 95% confidence intervals (CIs), and *P-*values. Because these outcomes were binary, treatment effects are expressed as odds ratios (ORs),and absolute event rates were presented as n/N (%). As a post-hoc sensitivity analysis, generalized linear mixed models (GLMMs) with a binomial distribution and logit link were fitted for these repeated binary outcomes to assess whether the direction and magnitude of treatment effects were consistent with the primary GEE analyses.

Neonatal Apgar scores were analyzed descriptively because of limited variability and a potential ceiling effect. Apgar scores are presented as median [interquartile range] and counts (percentages). Between-group comparisons of Apgar scores were performed using the Mann–Whitney U test, and categorical classifications were compared using Fisher’s exact test when applicable.

The Ramsay outcome was analyzed as a single binary outcome at the skin incision period(time = 5). Ramsay scores of 2–4 were coded as target sedation, and other scores were coded as non-target sedation. This outcome was analyzed using logistic regression and summarized using an odds ratio with a 95% confidence interval,and absolute event rates were presented as n/N (%).All statistical tests were two-sided, and a *P*-value < 0.05 was considered statistically significant.

Safety-related continuous outcomes, including systolic blood pressure, heart rate, and SpO_2_, were analyzed using the longitudinal models described above, and all valid measured values from participants included in the analysis were used. The Ramsay target-sedation outcome was analyzed using logistic regression, and Apgar scores were summarized descriptively with between-group comparisons when applicable.

To control the overall Type I error rate across multiple secondary outcomes evaluated via hypothesis testing (heart rate, SpO_2_,hurt, and anesthesia satisfaction), the Benjamini-Hochberg False Discovery Rate procedure and the conservative Bonferroni correction were prespecified for multiple comparisons. Both unadjusted and adjusted *P*-values are reported, with adjusted *P* < 0.05 defined as statistically significant.

Variables evaluated solely through descriptive statistics (Apgar scores) or presenting single-time-point exploratory evaluation (Ramsay Score)were not subjected to multiplicity adjustments.

Prespecified adverse clinical events, rescue interventions, and withdrawals due to safety criteria, if present, were summarized using counts and percentages.

All analyses were performed according to the assigned treatment group. The final analysis included all participants who completed the study procedures and had available outcome data. No participant met the prespecified withdrawal criteria requiring exclusion from the analysis.All participants included in the final analysis had available data for the outcome,no imputation was performed in this dataset.

## Main Results

### Baseline characteristics

A total of 55 parturients were included in the final analysis. 24 participants were randomly assigned to the normal saline group (NS group), received the intended intervention, and were analyzed for the outcome. 31 participants were randomly assigned to the esketamine group (KE group), received the intended intervention, and were analyzed for the outcome. The difference in group sizes arose by chance under unrestricted simple randomization.No participants were withdrawn, excluded, or lost to follow-up after randomization.The trial was not stopped early for safety, efficacy, or futility reasons. Recruitment ended at the end of the approved study period. The final analyzed sample was smaller than the feasibility-based target enrollment because emergency conversion from labor epidural analgesia to Category I cesarean section is uncommon, highly time-sensitive, and difficult to predict. In addition, the marked decline in birth rates in China since 2020 reduced the overall obstetric case volume, including at our center, which further limited the number of eligible parturients during the recruitment period. The participant screening, randomization, intervention, and analysis process is shown in Figure 1.

Baseline demographic characteristics and physiological variables were generally comparable between the two groups. No clear between-group differences were observed in age, body weight, baseline systolic blood pressure, heart rate, or SpO_2_. The distribution of baseline SpO_2_ values was also similar between groups. Details are shown in Table 1.

**Table 1.**
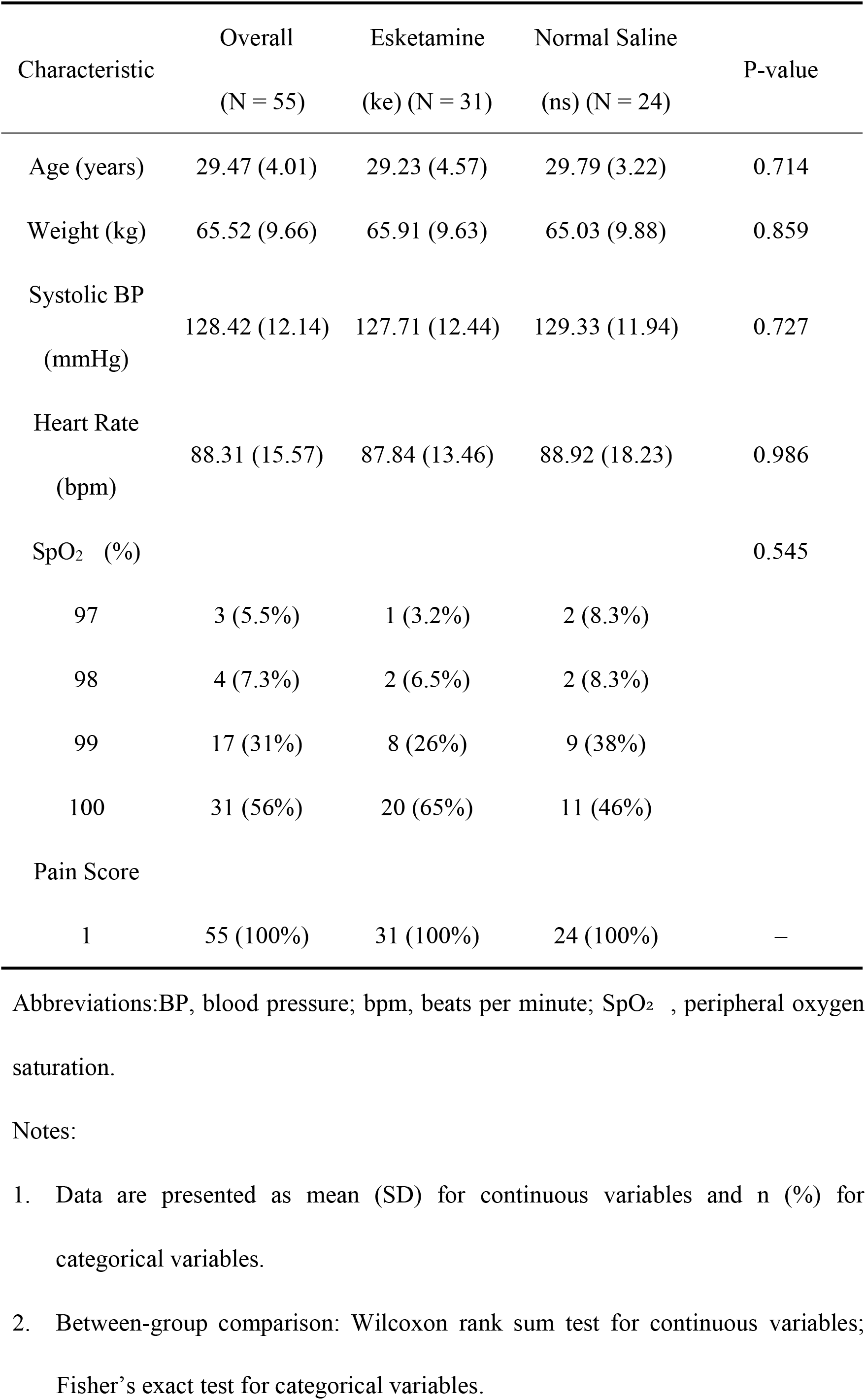
Patient Demographic and Baseline Characteristics.

| Characteristic | Overall<br>(N = 55) | Esketamine<br>(ke) (N = 31) | Normal Saline<br>(ns) (N = 24) | P-value |
| --- | --- | --- | --- | --- |
| Age (years) | 29.47 (4.01) | 29.23 (4.57) | 29.79 (3.22) | 0.714 |
| Weight (kg) | 65.52 (9.66) | 65.91 (9.63) | 65.03 (9.88) | 0.859 |
| Systolic BP<br>(mmHg) | 128.42 (12.14) | 127.71 (12.44) | 129.33 (11.94) | 0.727 |
| Heart Rate<br>(bpm) | 88.31 (15.57) | 87.84 (13.46) | 88.92 (18.23) | 0.986 |
| SpO<sub>2</sub> (%) |  |  |  | 0.545 |
| 97 | 3 (5.5%) | 1 (3.2%) | 2 (8.3%) |  |
| 98 | 4 (7.3%) | 2 (6.5%) | 2 (8.3%) |  |
| 99 | 17 (31%) | 8 (26%) | 9 (38%) |  |
| 100 | 31 (56%) | 20 (65%) | 11 (46%) |  |
| Pain Score |  |  |  |  |
| 1 | 55 (100%) | 31 (100%) | 24 (100%) | – |

Abbreviations:BP, blood pressure; bpm, beats per minute; SpO<sub>2</sub> , peripheral oxygen saturation.
Notes:
1. Data are presented as mean (SD) for continuous variables and n (%) for categorical variables.
2. Between-group comparison: Wilcoxon rank sum test for continuous variables;
Fisher's exact test for categorical variables.

Concomitant care was the same in both groups. All participants received routine perioperative monitoring, confirmation of epidural catheter patency, epidural alkalinized lidocaine according to the study protocol, and left the operating table after epidural administration to reduce supine hypotensive syndrome. No group-specific concomitant care was provided apart from the assigned study drug.The intervention and comparator were administered as intended in all participants. All 31 participants in the KE group received esketamine 0.15 mg/kg, and all 24 participants in the NS group received an equivalent weight-adjusted volume of normal saline 1 min before skin incision. No deviations from the assigned intervention or comparator occurred.No participant required vasopressor or chronotropic rescue treatment, conversion to general anesthesia, repeat neuraxial puncture, or withdrawal because of clinically significant hypotension or bradycardia during the observation period (20 min).Recruitment was conducted from [Sep 2023] to [May 2026]. Written informed consent was obtained from all participants.Maternal outcomes were followed for 20 min after study drug administration. Neonatal follow-up for the prespecified outcome was limited to Apgar assessment at 1, 5, and 10 min after birth.

### Primary outcome, systolic blood pressure

As illustrated in Figure 2, blood pressure in the esketamine group demonstrated a transient upward trend during the early period from 0 to 5 minutes, whereas systolic blood pressure(SBP)in the normal saline group remained relatively stable over the same interval. Thereafter, both groups showed a mild downward trend over time. In addition, the spaghetti plot revealed substantial inter-individual variability in SBP trajectories despite generally similar group-level temporal patterns.

**Figure 2.**
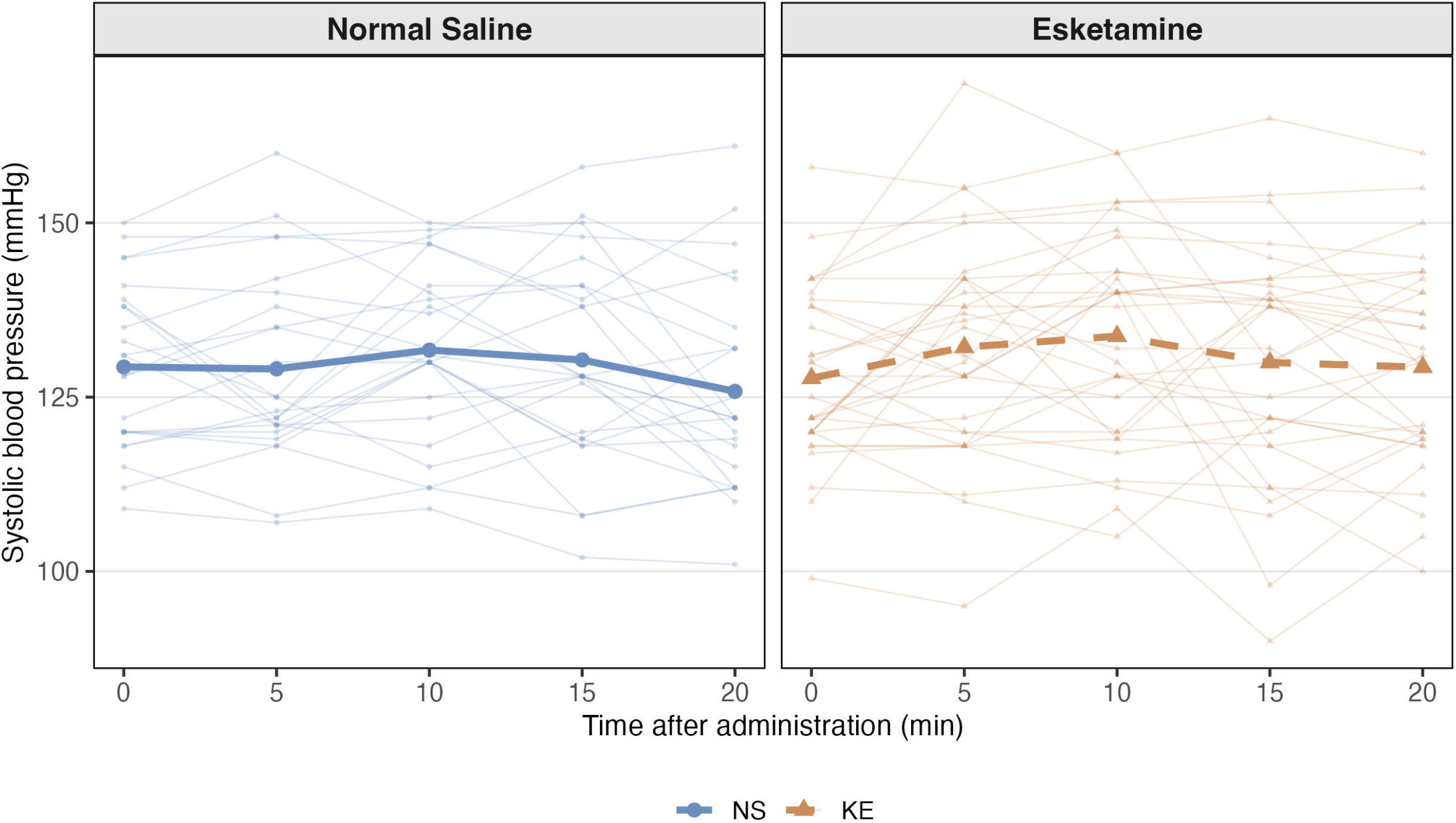
Systolic blood pressure trajectories over time

To evaluate longitudinal changes in systolic blood pressure (SBP), we fitted a linear mixed-effects model with random intercepts for individual participants to account for within-subject correlation from repeated measurements (Table 2). We formally evaluated both linear and non-linear polynomial time terms; although the inclusion of a quadratic time term showed a marginally statistical significant improvement in model fit over the linear-only specification (AIC = 1637.1 vs. 1635.8; Likelihood Ratio Test *P* = 0.070), the quadratic model was retained as our primary specification because it better reflects the clinical physiology of an initial SBP elevation followed by a decline. Model diagnostics confirmed that the conditional residuals closely conformed to Gaussian normal distributions on Q-Q plots. Furthermore, linear specification models were evaluated as a sensitivity analysis, yielding completely consistent treatment effect estimates.

**Table 2.**
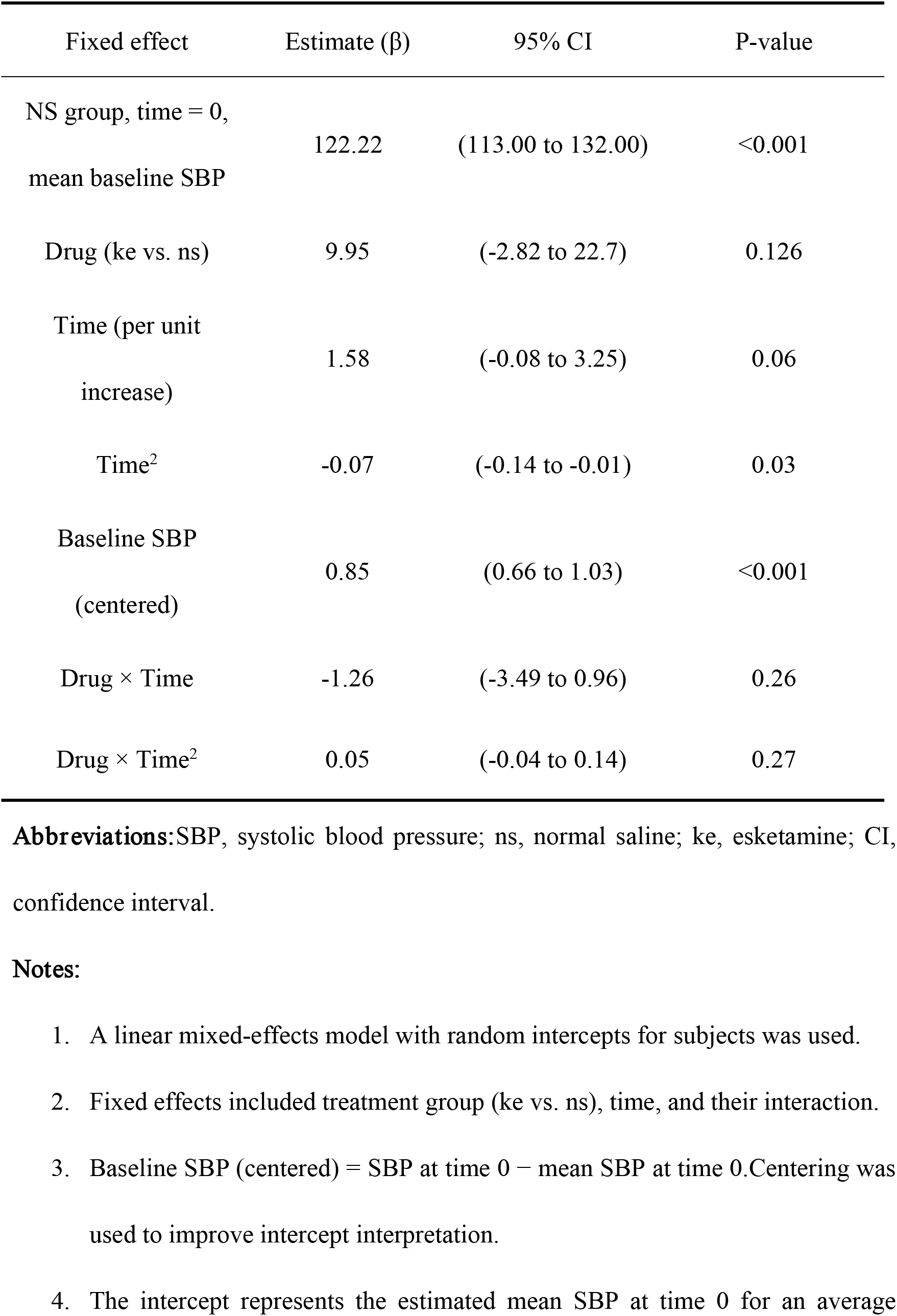
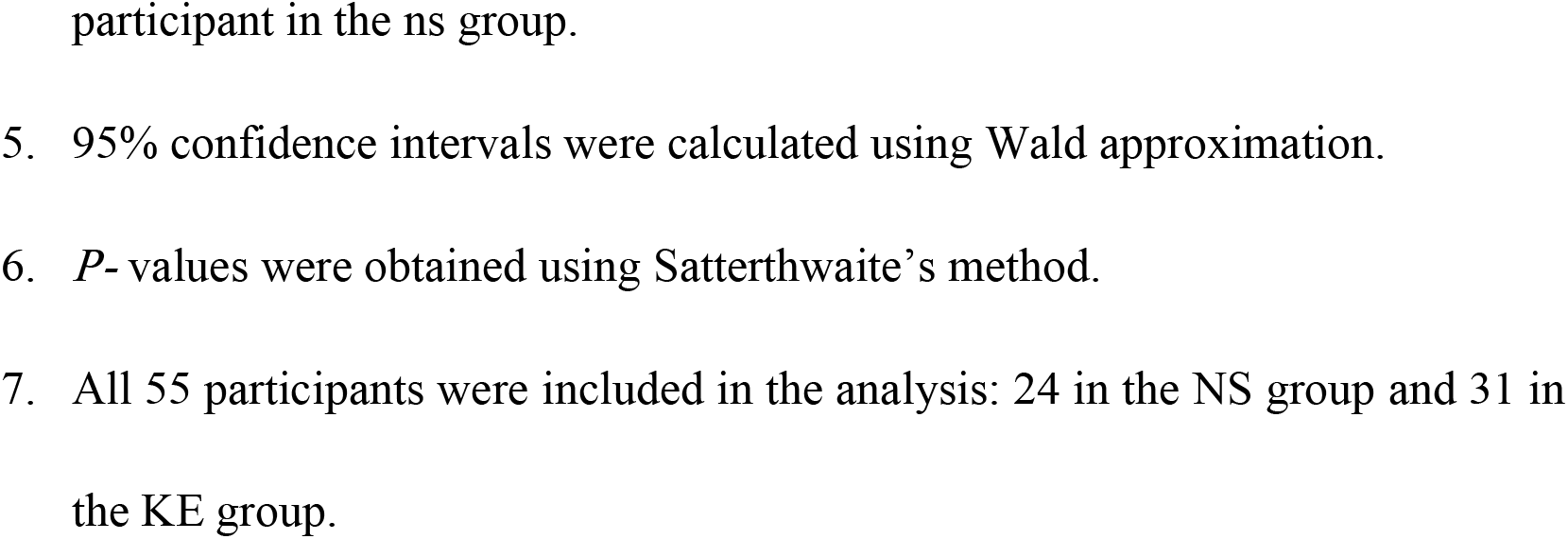
Linear Mixed-Effects Model for Longitudinal Changes in Systolic Blood Pressure (SBP) with Centered Baseline Adjustment.

In our primary quadratic model, overall SBP levels did not differ statistical significantly between the esketamine and normal saline groups (*β* = 9.95, 95% CI: -2.82 to 22.72, *P* = 0.126). Regarding temporal dynamics, while the linear time component showed a non-statistical significant increasing trend (*β* = 1.58, 95% CI: -0.08 to 3.25, *P* = 0.062), the quadratic time term was statistically significant (*β* = -0.07, 95% CI: -0.14 to -0.01, *P* = 0.030), capturing the non-linear trajectory over time. However, neither the linear group-by-time interaction (*β* = -1.26, 95% CI: -3.49 to 0.96, *P* = 0.264) nor the quadratic group-by-time interaction (*β* = 0.05, 95% CI: -0.04 to 0.14, *P* = 0.271) reached statistical significance, indicating no substantial difference in SBP trajectories between the two groups.

Centered baseline SBP remained a strong independent predictor of subsequent SBP measurements (*β* = 0.85, 95% CI: 0.66 to 1.03, *P* < 0.001), showing that participants with higher initial SBP relative to the cohort mean consistently maintained higher values throughout the observation period. Under this parameterization, the model intercept indicated an estimated mean SBP of 122.22 mmHg (95% CI: 113.00 to 132.00, *P* < 0.001) at time zero for an average participant in the normal saline group.

### continuous secondary outcomes:heart rate and SpO_2_

As illustrated in Figure 3, the mean heart rate at time 0 was comparable between the two groups. Following drug administration, the first post-administration measurement at time 5 minutes showed a transient increase in heart rate in the esketamine group, whereas the normal saline group showed a slight decrease over the same interval.

**Figure 3.**
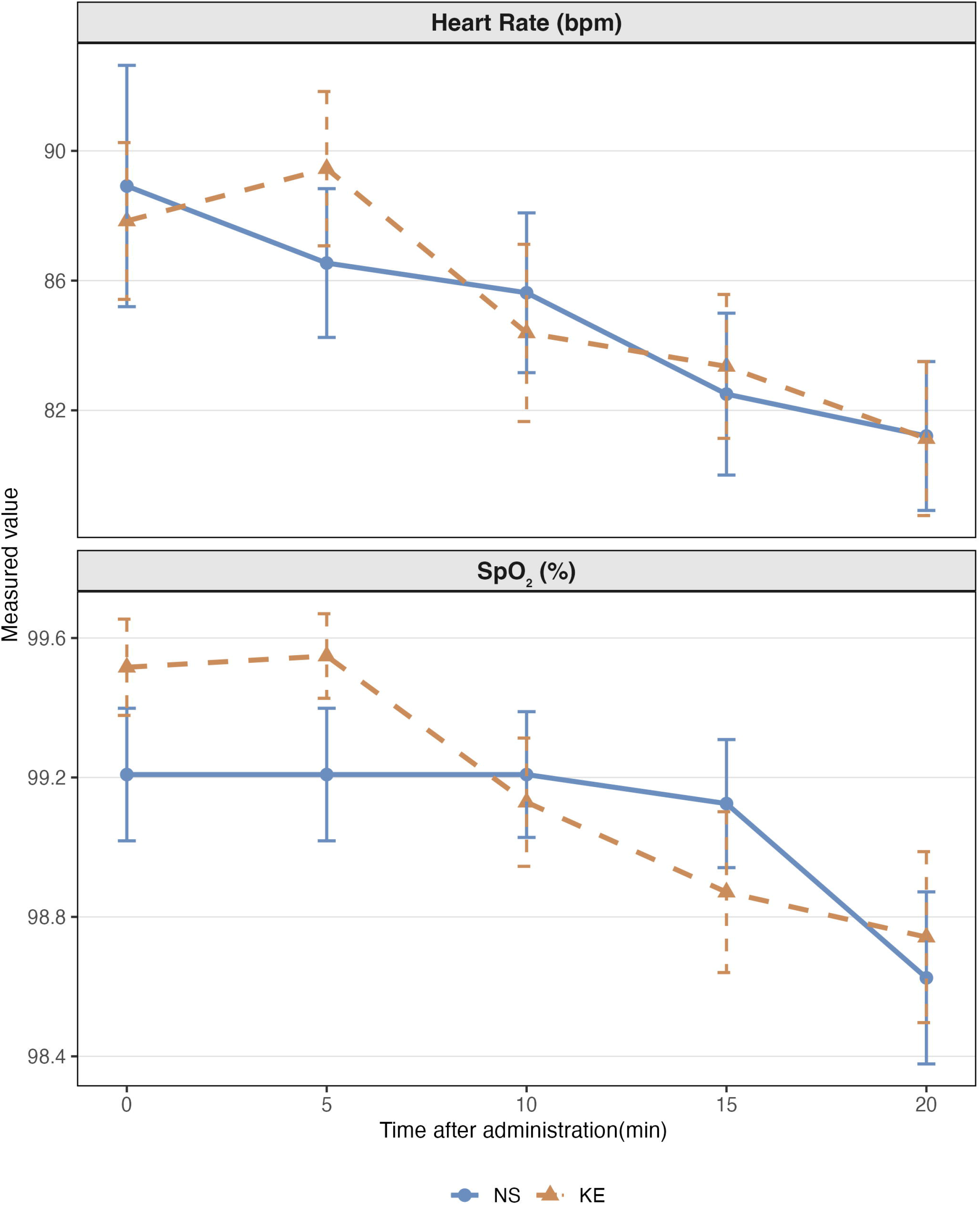
Heart rate and SpO2 trajectories over time

Thereafter, the mean heart rate in both groups gradually declined over time.

To evaluate longitudinal changes in heart rate (HR), we applied the same linear mixed-effects model framework as that used for systolic blood pressure (Table 3). Model comparison between linear and quadratic time specifications showed that the inclusion of a quadratic time term did not improve model fit (AIC = 1664.1 vs 1661.1;Likelihood Ratio Test *P =* 0.655).Following the principle of parsimony, the standard linear mixed-effects model was retained as our primary specification. Visual inspection of conditional residual Q-Q plots confirmed that the model assumptions closely conformed to Gaussian normal distributions.The primary model included treatment group, continuous time, the group-by-time interaction, and centered baseline HR as fixed effects, alongside random intercepts for individual participants to account for within-subject correlation from repeated measurements. Baseline HR was centered at the sample mean prior to analysis to enhance intercept interpretability.

**Table 3.**
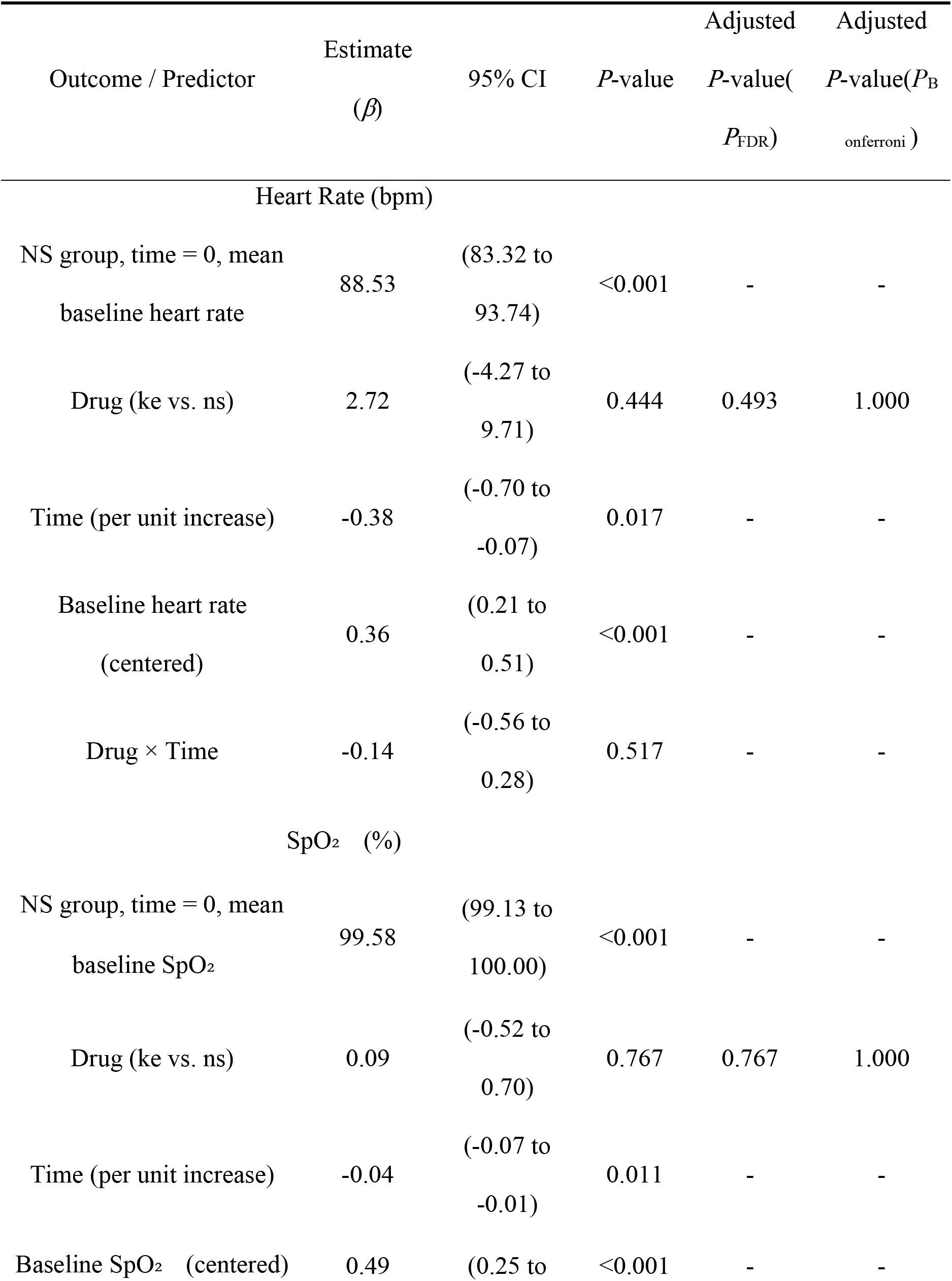
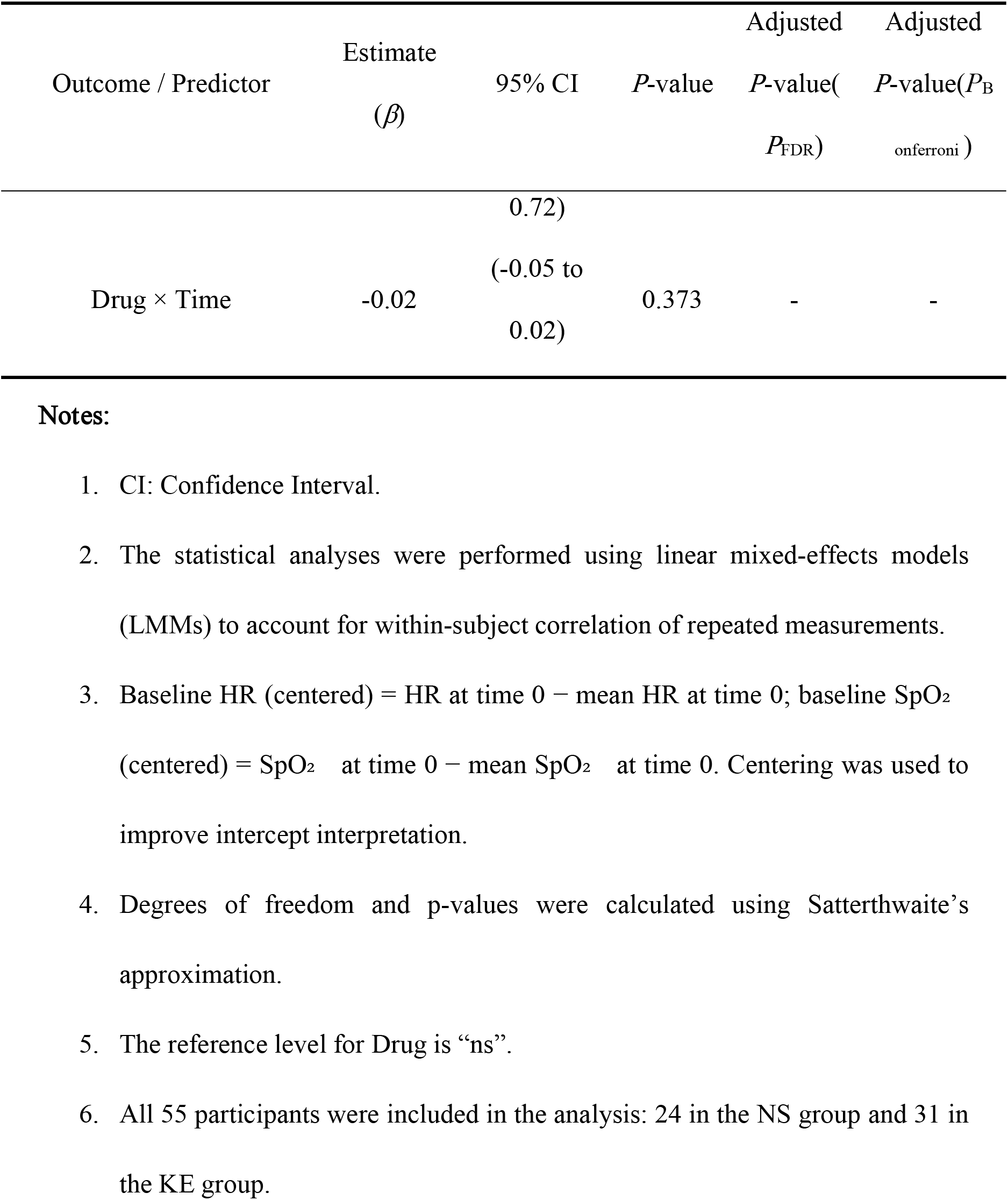
Linear Mixed Effects Model Results for Secondary Outcomes (Heart Rate and SpO₂)-.

We can’t observe significant main effect in treatment group(*β*=2.72, *P*= 0.444,95%CI:-4.27 to 9.71), and the group-by-time interaction was not statistically significant (*β*=−0.14, *P*=0.517,95%CI:-0.56 to 0.28). A modest but statistically significant decrease in heart rate over time was identified overall (*β*=−0.38, *P* = 0.017,95%CI:-0.70 to -0.07). We can observe that centered baseline heart rate is strongly associat with subsequent heart rate measurements (*P* < 0.001), indicating that participants with higher initial heart rate tended to maintain higher heart rate throughout the follow-up period,these findings also suggest that intravenous esketamine at the administered dose did not produce clinically meaningful alterations in perioperative heart rate compared with normal saline.

As illustrated in Figure 3, mean oxygen saturation (SpO_2_) in the esketamine group was slightly higher than that in the normal saline group at time 0, and a similar between-group difference was generally maintained throughout the observation period. Following drug administration, we can observe that SpO_2_ gradually decrease in both groups over time, with a visually steeper decline in the normal saline group compared with the esketamine group.

To evaluate longitudinal changes in oxygen saturation (SpO_2_), we applied the same linear mixed-effects framework as used for systolic blood pressure (Table 4). Model comparison between linear and quadratic time specifications indicated that adding a quadratic time term did not yield a statistically significant improvement in model fit (AIC = 591.16 vs. 590.80; Likelihood Ratio Test *P* = 0.1618). Based on the principle of parsimony, the linear mixed-effects model was selected as our primary specification.Visual inspection of conditional residual Q-Q plots revealed that while central residuals conformed well to Gaussian normality, mild left-tail deviations were present due to the boundary and ceiling effects inherent to SpO_2_ measurements.

**Table 4.**
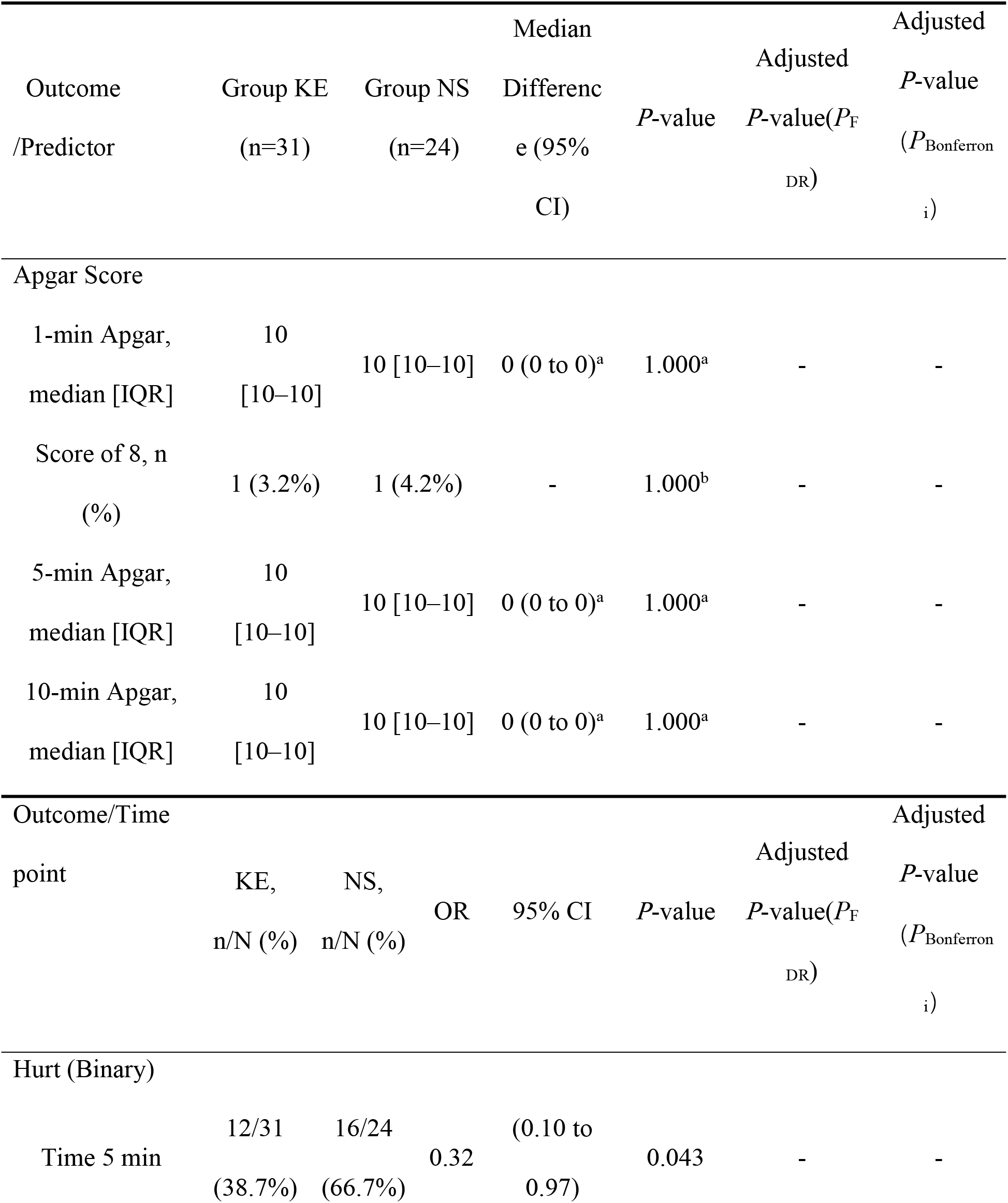
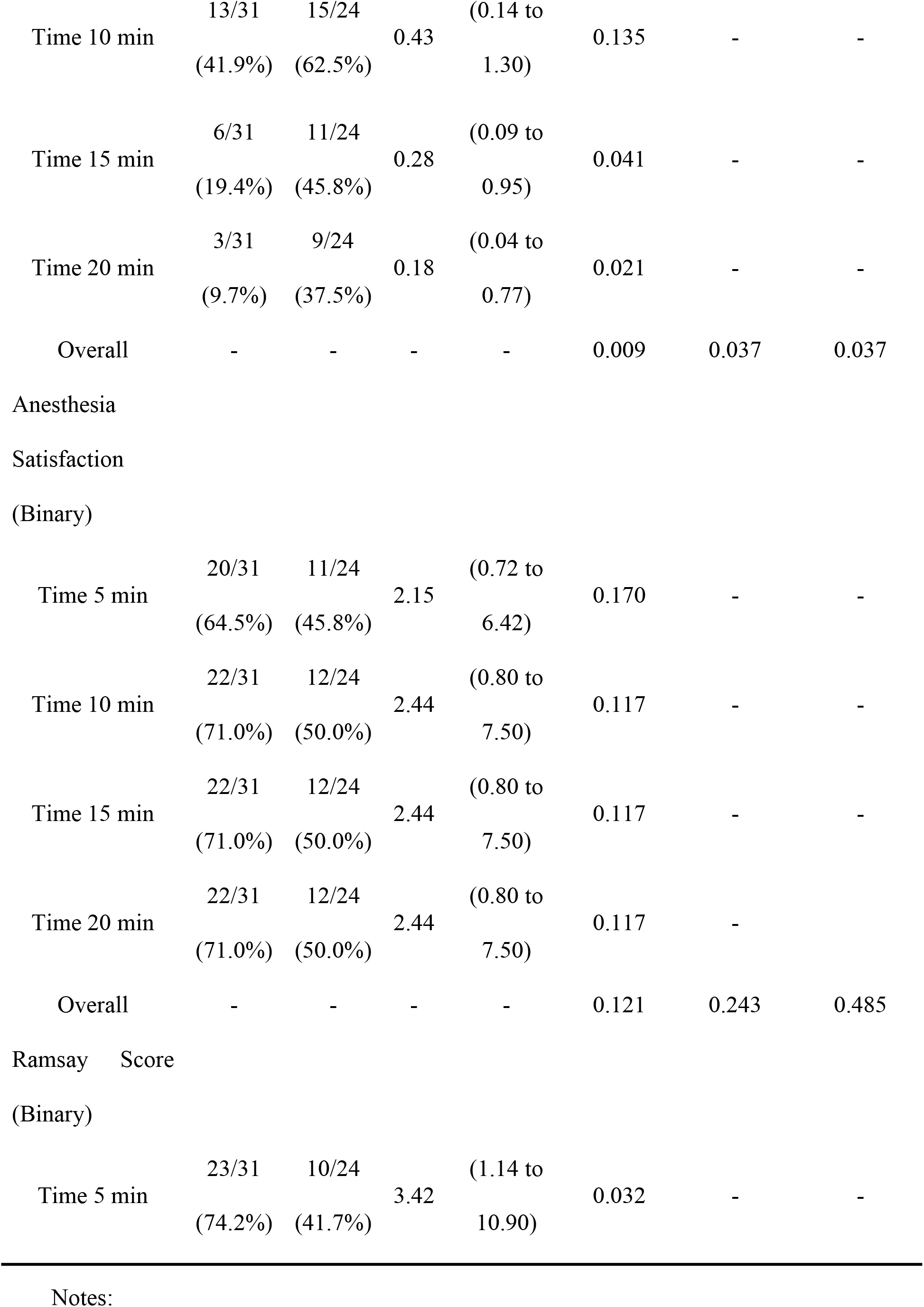
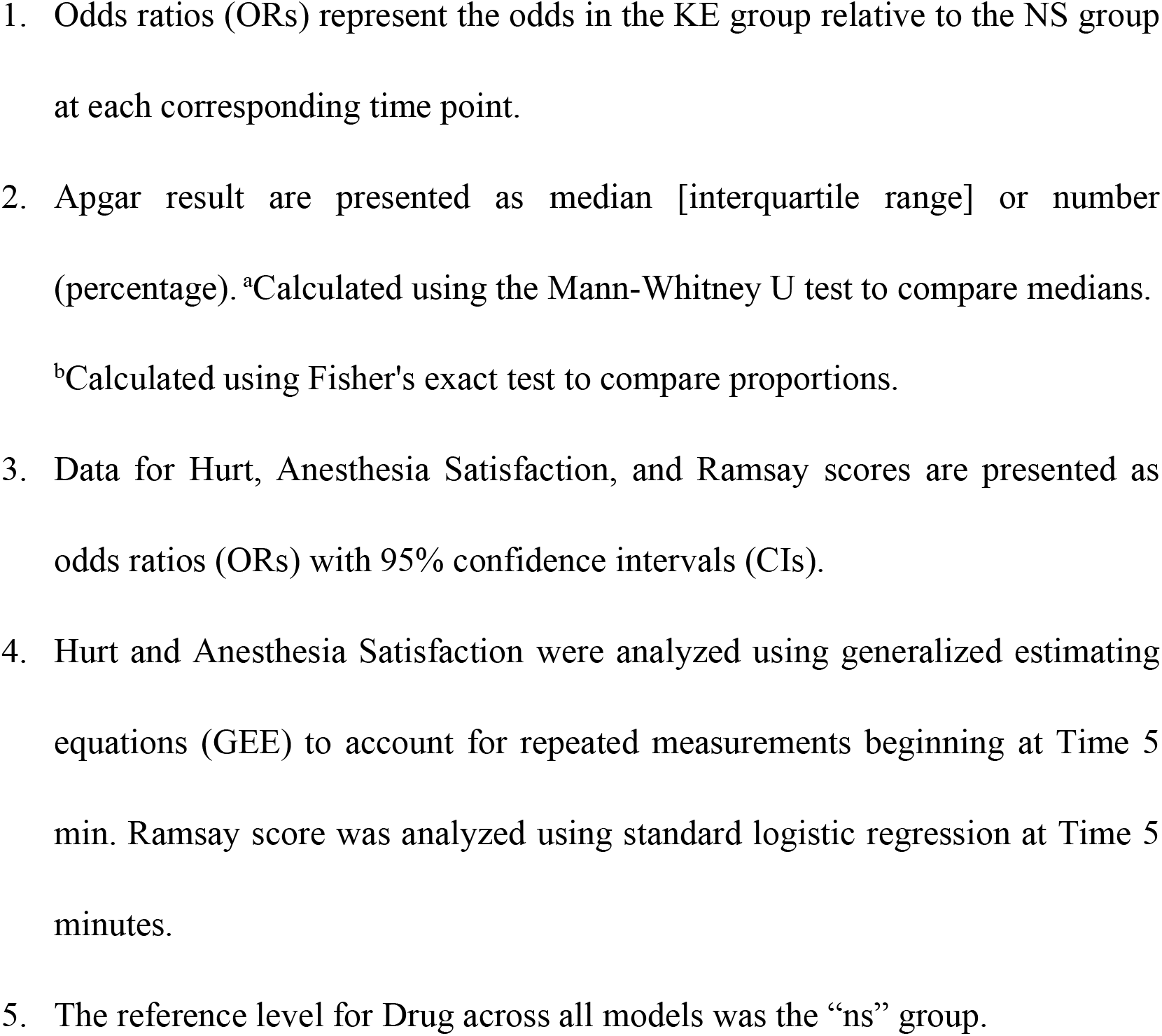
Multivariable analysis of secondary outcomes: Apgar score, Hurt, Satisfaction, and Ramsay score.

We can’t observe significant main effect of treatment group on SpO_2_ (*β*=0.09, *P*= 0.767,95%CI:-0.52 to 0.70), indicating similar oxygen saturation profiles between the esketamine and normal saline groups during the study period. Likewise, no significant group-by-time interaction was observed (*β*=−0.02, *P* = 0.373,95%CI:-0.05 to 0.02), suggesting that the longitudinal pattern of SpO_2_ change was comparable between groups.Overall, SpO_2_ remained clinically stable across both groups, and no evidence suggested that esketamine adversely affected perioperative oxygenation. While the model indicated a slight but statistically significant decline in SpO_2_ over time (*β*= -0.04, *P* = 0.011,95%CI:-0.07 to -0.11), this trend was consistent across both groups, with no drug-by-time interaction. Centered baseline SpO_2_ was strongly associated with subsequent measurements (*P*< 0.001), indicating that participants with higher initial oxygen levels tended to maintain better oxygenation throughout the follow-up period.

Sensitivity analysis using a beta generalized linear mixed-effects model (GLMM) further confirmed the robustness of these findings(See the Supporting Information S2 Table). Consistent with the primary LMM analysis, the beta model revealed no significant between-group differences at any time point, including 5 min (OR = 1.35, 95% CI: 0.86 to 2.12, *P* = 0.187), 10 min (OR = 0.91, 95% CI: 0.59 to 1.40, *P* = 0.666), 15 min (OR = 0.80, 95% CI: 0.53 to 1.21, *P* = 0.282), and 20 min (OR = 0.99, 95% CI: 0.66 to 1.48, *P* = 0.945).

### binary secondary outcomes:hurt,anesthesia satisfaction and ramsay score

Figure 4 illustrates the temporal trend of intraoperative hurt in both groups over time. At time 0, patients in both groups generally reported pain; however, the incidence of hurt gradually decreased during the observation period. Compared with the normal saline group, the esketamine group demonstrated a more pronounced decline in pain, particularly during the early phase following administration (0-5 min). Over time, the proportion of patients reporting no pain progressively increased in the esketamine group, whereas the normal saline group maintained a relatively higher proportion of pain reports throughout the study period. Visually, esketamine appeared to provide consistently better intraoperative analgesia across most time points.

**Figure 4.**
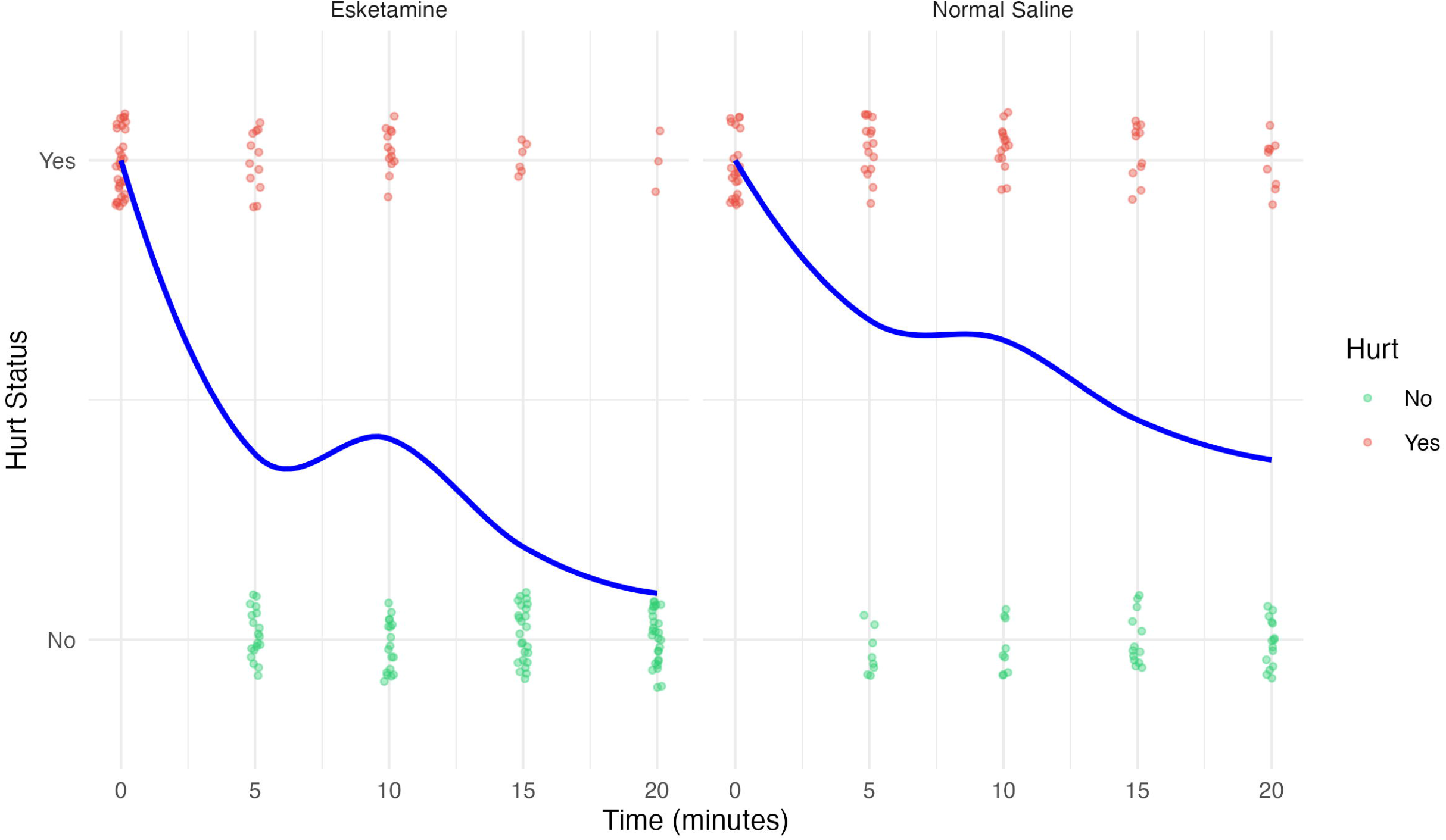
Intraoperative pain over time by group

We further analyzed hurt as a repeated binary outcome using generalized estimating equations (GEE) with a logit link function in the geepack::geeglm package (Table 4). We used an autoregressive correlation structure [AR(1)] to account for within-subject correlations across repeated observations. Robust sandwich variance estimators provided standard errors, and we derived 95% CIs and *P*-values from these estimates. We present the results as odds ratios (ORs) with 95% CIs, together with the observed n/N and percentages at each time point.During the 20-min observation period, fewer participants reported hurt in the esketamine group than in the normal saline group at most time points. At 5 min, 12/31 participants (38.7%) in the esketamine group reported hurt, compared with 16/24 participants (66.7%) in the normal saline group. The esketamine group had significantly lower odds of hurt at this time point (OR = 0.32, 95% CI: 0.10 to 0.97, *P* = 0.043).At 10 min, 13/31 participants (41.9%) in the esketamine group and 15/24 participants (62.5%) in the normal saline group reported hurt. Although the proportion remained lower in the esketamine group, the between-group difference did not reach statistical significance (OR = 0.43, 95% CI: 0.14 to 1.30, *P* = 0.135).At 15 min, 6/31 participants (19.4%) in the esketamine group reported hurt, compared with 11/24 participants (45.8%) in the normal saline group.

The esketamine group again showed significantly lower odds of hurt (OR = 0.28, 95% CI: 0.09 to 0.95, *P* = 0.041). At 20 min, 3/31 participants (9.7%) in the esketamine group and 9/24 participants (37.5%) in the normal saline group reported hurt, with significantly lower odds in the esketamine group (OR = 0.18, 95% CI: 0.04 to 0.77, *P* = 0.021).Sensitivity analyses using generalized linear mixed models (GLMMs) yielded similar findings, supporting the robustness of the primary GEE results.

We further analyzed anesthesia satisfaction as a repeated binary outcome using generalized estimating equations (GEE) with a logit link function in the geepack::geeglm package (Table 4). We used an autoregressive correlation structure [AR(1)] to account for within-subject correlations across repeated observations.

Robust sandwich variance estimators provided standard errors, and we derived 95% CIs and *P*-values from these estimates. We present the results as odds ratios (ORs) with 95% CIs, together with the observed n/N and percentages at each time point.

During the 20-min observation period, the esketamine group showed a higher proportion of favorable anesthesia satisfaction than the normal saline group at all evaluated time points. At 5 min, 20/31 participants (64.5%) in the esketamine group had favorable anesthesia satisfaction, compared with 11/24 participants (45.8%) in the normal saline group. The odds ratio was 2.15, but the between-group difference did not reach statistical significance (95% CI: 0.72 to 6.42, *P* = 0.170).At 10 min, favorable anesthesia satisfaction occurred in 22/31 participants (71.0%) in the esketamine group and 12/24 participants (50.0%) in the normal saline group. The odds ratio was 2.44, with a 95% CI crossing unity (95% CI: 0.80 to 7.50, *P* = 0.117). The same pattern appeared at 15 and 20 min, where 22/31 participants (71.0%) in the esketamine group and 12/24 participants (50.0%) in the normal saline group had favorable anesthesia satisfaction at both time points. The corresponding odds ratios remained 2.44 at 15 min and 20 min, respectively, with identical 95% CIs and *P*-values (95% CI: 0.80 to 7.50, *P* = 0.117 for both).Sensitivity analyses using generalized linear mixed models (GLMMs) yielded similar findings, supporting the robustness of the primary GEE results.

We analyzed the Ramsay score at the first post-administration assessment as a single-measurement binary outcome using logistic regression. We defined target sedation as a Ramsay score of 2 – 4 and reported the outcome as n/N (%). Target sedation occurred more frequently in the esketamine group than in the normal saline group [23/31 (74.2%) vs. 10/24 (41.7%)]. The esketamine group showed significantly higher odds of achieving target sedation than the normal saline group (OR = 3.42, 95% CI: 1.14 to 10.90, *P* = 0.032).

### neonatal outcome

Apgar scores remained uniformly high in both groups(Table 4). At the 1-min assessment, one neonate in each group had an Apgar score of 8, while all remaining neonates had a score of 10. At the 5- and 10-min assessments, all neonates achieved Apgar scores of 10. Consequently, there were no statistically significant differences in Apgar scores between the groups at any time point (all median differences = 0, 95% CI: 0 to 0; all *P* = 1.000). These descriptive findings did not suggest an adverse effect of esketamine on early neonatal condition.

### Multiple Comparison Adjustments Across Secondary Outcomes

The adjusted *P* values were consistent with the conclusions based on the unadjusted analyses. Among the secondary outcomes, Hurt demonstrated a statistically significant difference (unadjusted *P* = 0.009). This difference remained statistically significant after adjustment for multiple comparisons using both the Benjamini – Hochberg false discovery rate procedure (*P*_FDR_ = 0.037) and the conservative Bonferroni method (*P*_Bonferroni_ = 0.037). Heart rate (unadjusted *P* =0.444,*P*_FDR_ = 0.592; *P*_Bonferroni_ = 1.000), SpO_2_ (unadjusted *P* =0.767,*P*_FDR_ = 0.767; *P*_Bonferroni_ = 1.000), and anesthesia satisfaction (unadjusted *P* =0.121,*P*_FDR_ = 0.242; *P*_Bonferroni_ = 0.485) showed no statistically significant between-group differences after adjustment.

## Discussion

The effects of low-dose esketamine were evaluated during the clinically critical 20-min window of Category I emergency cesarean section. The primary endpoint was the longitudinal change in systolic blood pressure (SBP). The mixed-effects model included quadratic time terms to describe the post-administration SBP trajectory. The model indicated quadratic patterns of change in SBP over time in both groups.

Although Figure 2 suggests a transient early increase in SBP in the esketamine group between 0 and 5 min, compared with a relatively stable pattern in the normal saline group, this visual tendency was not supported by the mixed-effects model. Thus, within the dose and observation window of this study, esketamine showed a limited overall effect on maternal circulation and did not produce a clear signal of hemodynamic instability. [11–12].Avoiding maternal hypotension is particularly relevant in obstetric anesthesia, as hypotension may reduce placental perfusion, impair fetal oxygenation, and increase the risk of fetal acidosis [19]. This concern is even more important in Category I emergency cesarean section, where fetal reserve may already be compromised and time for correction is limited [2,15]. In our analysis, esketamine was not associated with decrease in SBP.Instead, the fitted SBP values have a higher trajectory in the esketamine group at all four post-administration assessment times,although this change does not reach the statistically significant.At the same time,these nonsignificant findings provided no clear evidence that esketamine had an additional effect on maternal SBP fluctuations emergency cesarean section. Compared with many conventional intravenous sedative-analgesic agents, which may be accompanied by blood pressure reduction, low-dose esketamine may offer some advantage in maintaining maternal circulatory stability, [7,9]Baseline SBP was strongly associated with subsequent SBP measurements, indicating that the initial hemodynamic state continued to influence perioperative SBP trajectories in this highly stressful setting. The use of a subject-specific random intercept allowed each parturient to have her own baseline SBP level and accounted for within-subject correlation and between-subject heterogeneity. This was consistent with Figure 1, which showed marked individual variability despite relatively similar group-level trends. For this type of repeated continuous outcome, adjustment for baseline values together with random intercepts is therefore more informative than a simple comparison of group mean changes.Previous perioperative studies have reported broadly similar findings: at analgesic-sedative doses, ketamine or esketamine generally has limited overall circulatory effects [8,11–12]. However, transient early increases in blood pressure have also been described. In a retrospective study of esketamine use during emergency cesarean delivery, SBP increased significantly within 5 – 10 min after administration [20]. In our cohort, the early upward SBP tendency in the esketamine group did not remain significant after baseline adjustment and modeling of repeated measurements. The lower dose used here, the 5-min measurement interval, and the exclusion of patients with hypertension or overt cardiovascular disease may all have reduced the likelihood of detecting a brief pressor response. In addition, hemodynamics during Category I cesarean section are influenced by multiple perioperative factors, including fetal delivery, uterine manipulation, oxytocin administration, and rapidly changing stress responses [15].

For the continuous secondary outcomes, no significant treatment effect or treatment-by-time interaction was detected for heart rate or SpO_2_. Similar to the SBP model, baseline values and subject-specific random intercepts were included in the HR and SpO_2_ models. Baseline HR and baseline SpO_2_ were both strongly associated with subsequent measurements, again emphasizing the importance of individual physiological status during emergency obstetric anesthesia.Figure 2 showed a transient early increase in HR in the esketamine group, whereas HR in the normal saline group declined slightly. This apparent difference, however, was not statistically significant in the mixed-effects model. Both HR and SpO_2_ showed mild downward trends over time. These patterns are more likely to reflect shared perioperative physiological changes rather than a drug-specific adverse effect . As neuraxial anesthesia becomes more complete, pain and stress responses decline, which may reduce sympathetic activity [6]. Relief of aortocaval compression after fetal delivery, left uterine displacement, oxytocin use, and perioperative ventilatory changes may also contribute to alterations in HR and SpO_2_. Overall, these mild changes are more consistent with gradual perioperative stabilization than with esketamine-related circulatory or respiratory deterioration.Although the time effect for SpO_2_ reached statistical significance, values remained clinically acceptable throughout the observation period. There was no statistical significant between-group difference and no treatment-by-time interaction, indicating that esketamine did not produce detectable respiratory depression at the dose used in this study. This is consistent with pharmacological and clinical reviews suggesting that ketamine-class drugs, at analgesic-sedative doses, cause relatively limited respiratory depression and may preserve spontaneous respiration and upper airway reflexes [7–8]. No worsening trend in SpO_2_ was observed in the esketamine group [11–12].The relatively stable hemodynamic profile observed here may reflect the complex sympathomodulatory effects of esketamine. Ketamine-class drugs are traditionally associated with increased blood pressure and heart rate, effects generally attributed to sympathetic activation and inhibition of catecholamine reuptake [7,9,23]. In the perioperative setting, however, analgesia, relief of anxiety, and attenuation of the stress response may offset this sympathostimulatory effect, resulting in little net hemodynamic change [8,12]. Stereoselective cardiovascular pharmacodynamics may also contribute to the observed response [21]. In this study, the low dose, the exclusion of high-risk cardiovascular patients, and the 5-min measurement schedule may have further limited the detection of a short-lived pressor peak [11,20].

Among the binary outcomes, the time course of intraoperative pain favored esketamine during the 20-min observation period, but the analgesic pattern varied with the surgical stage. At 5 min, the assessment generally corresponded to skin incision and subcutaneous dissection. These steps mainly produce sharp somatic pain, and low-dose esketamine appeared to cover this early nociceptive stimulus effectively. In line with this clinical sequence, fewer patients in the esketamine group reported pain at 5 min than in the normal saline group.At 10 min, however, the between-group difference did not reach statistical significance, although the point estimate still favored esketamine. This finding is clinically understandable. In our operative workflow, the 10-min assessment often coincided with the most stimulating part of the cesarean delivery, including abdominal traction, fundal pressure to assist fetal delivery, fetal extraction, and manual removal of the placenta. These steps can produce intense somatic and visceral stimulation. A subanesthetic dose of esketamine may reduce discomfort, but it may not fully cover this peak surgical stimulation on its own, especially when the epidural block has not yet reached a complete surgical level.

At 15 and 20 min, the analgesic advantage of esketamine became more evident again. By this stage, the most intense operative steps had usually passed, and the epidural local anesthetic had more time to reach an adequate surgical level. Therefore, the lower occurrence of pain in the esketamine group at these later time points likely reflects the combined effect of esketamine and the progressively established neuraxial block. This time-dependent pattern supports the role of low-dose esketamine as a bridging analgesic during the vulnerable interval before epidural conversion becomes fully effective, rather than as a substitute for adequate neuraxial anesthesia [3,8,12].

These results are consistent with previous studies of low-dose ketamine or esketamine during cesarean section under neuraxial anesthesia. Traction, peritoneal stimulation, uterine manipulation, and fundal pressure can cause visceral pain that local anesthetics may not fully cover. In this setting, additional intravenous analgesia or sedation can be clinically useful. Previous studies have shown that low-dose esketamine can reduce intraoperative pain scores, decrease the need for rescue analgesia, and improve perioperative comfort [8,11,12]. Our findings extend this evidence to Category I emergency cesarean section and highlight a practical point: low-dose esketamine may work best as a short-term bridge for early and moderate surgical stimulation, while its effect may be insufficient during the most intense moments of delivery.

Esketamine also increased the probability of achieving the target Ramsay sedation level, defined as scores of 2, 3, or 4. This suggests that low-dose esketamine can provide clinically acceptable perioperative sedation [7–8]. During cesarean section under neuraxial anesthesia, moderate sedation may reduce anxiety, improve comfort, and facilitate cooperation without requiring deep sedation. In this cohort, sedation remained within an acceptable range, with no obvious oversedation, respiratory depression, or clinically meaningful deterioration in SpO_2_. These findings suggest a favorable balance between analgesia, sedation, and maternal safety in this emergency setting .By contrast, surgeon satisfaction with muscle relaxation did not differ significantly between groups at any time point, although the point estimates tended to favor esketamine. This suggests that esketamine mainly improves maternal comfort and the analgesic-sedative experience, rather than correcting insufficient motor block or replacing adequate neuraxial anesthesia. It should therefore be viewed as a bridging adjunct, not as a definitive solution to failed or incomplete epidural conversion [3,8]. This interpretation is consistent with prior work showing that low-dose esketamine mainly improves analgesia and sedation, with limited direct effect on muscle relaxation [11–12].The analgesic and sedative effects of esketamine are pharmacologically plausible. Esketamine is the S-enantiomer of ketamine and has higher affinity for N-methyl-D-aspartate (NMDA) receptors, allowing more pronounced analgesic and dissociative sedative effects at lower doses [10,23]. NMDA receptor antagonism may reduce central sensitization and amplification of visceral pain, thereby attenuating discomfort from traction and peritoneal stimulation. Its dissociative sedative properties may also reduce anxiety, fear, and perioperative stress, improving subjective comfort and cooperation [9]. These features may explain why low-dose esketamine is particularly suitable for a brief bridging role in Category I emergency cesarean section [7–8,11–12].

Also there is no difference in Apgar score was observed between groups. Apgar scores followed a similar pattern over time between groups, and there was no esketamine-related worsening trend. This finding is consistent with previous reports suggesting that low-dose ketamine-class agents used during cesarean section do not usually produce clear adverse effects on early neonatal Apgar scores [11–12].Although esketamine can cross the placenta, neonatal condition in this setting may depend less on drug exposure alone and more on the stability of maternal hemodynamics and oxygenation. Low-dose ketamine-class drugs may preserve maternal circulation and spontaneous respiration better than sedatives associated with hypotension or respiratory depression, which could theoretically help maintain placental perfusion and fetal oxygen supply [7–8,12]. In this study, no obvious hypotension, severe hypoxemia, or worsening of Apgar scores was observed, indirectly supporting this interpretation.

These findings are clinically relevant because the study focused on a narrow and difficult window in obstetric anesthesia: emergency conversion from labor analgesia to Category I cesarean section before neuraxial blockade has become fully established. Beyond the extreme time pressure, failed conversion of labor epidural analgesia to surgical anesthesia remains an important concern. Although low-concentration local anesthetics may provide satisfactory labor analgesia, subsequent epidural top-up with high-concentration local anesthetics, such as 2% lidocaine, can still result in incomplete, unstable, or patchy block, with reported failure rates ranging from 0% to 21% [3,30]. Contributing factors may include suboptimal epidural catheter position, prolonged low-concentration local anesthetic infusion, catheter migration during labor, and insufficient time for high-concentration local anesthetics to spread adequately in Category I emergency cesarean section [3]. Consequently, some patients may still experience significant intraoperative pain despite having an indwelling epidural catheter [30].In this context, the purpose of the present study was not to replace adequate neuraxial anesthesia, but to examine whether low-dose intravenous esketamine could serve as a short-term bridging adjunct during the interval before a sufficient surgical block is achieved. The observed reduction in intraoperative hurt and the higher probability of target sedation suggest that esketamine may provide clinically useful analgesic-sedative support in this brief but high-risk period, without adding a clear circulatory, respiratory, or neonatal safety signal [3,11–12,30].When Category I emergency cesarean section is decided, transport safety also becomes an important practical issue. To achieve a surgical anesthetic level as quickly as possible, some centers initiate epidural administration of high-concentration local anesthetics in the labor room or during transfer to the operating room. A 2008 survey of 209 hospitals in the United Kingdom reported that 68% completed the full epidural top-up in the labor room, and most units did not use monitoring during transport [25]. This practice may save time, but potential risks include local anesthetic systemic toxicity, severe allergic reactions, excessively high block, and supine hypotensive syndrome. Without continuous vital sign monitoring, fetal heart rate monitoring, rescue drugs, and full resuscitation capability, such practice represents a trade-off between time efficiency and perioperative safety [26].Our center does not initiate epidural drug administration during transfer. Instead, dosing is performed after arrival in the operating room under routine monitoring. This approach further shortens the time available for establishing a surgical block, but it allows earlier recognition and management of adverse events under more complete monitoring and resuscitation conditions. Within this more conservative management model, low-dose intravenous esketamine did not show a clear adverse circulatory or respiratory signal.During conversion from labor analgesia to cesarean section, adding sodium bicarbonate to lidocaine is a commonly studied rapid-onset strategy. By altering the physicochemical properties of lidocaine, alkalinization can shorten the time required to establish an adequate surgical anesthetic level, which is particularly relevant in obstetric emergencies [4,27–28]. However, drug compatibility and precipitation after preparation remain important safety considerations. At our center, labor epidural analgesia is maintained with 0.09% ropivacaine, and alkalinized lidocaine was used during emergency conversion to Category I cesarean section to accelerate epidural onset. A case report and in vitro study from Peking Union Medical College Hospital already proved that adding alkalinized lidocaine after routine 0.09% ropivacaine labor analgesia was clinically acceptable [29].Another strength of the present study is its distinctive clinical context. Many previous studies evaluating intravenous esketamine during cesarean delivery have focused on elective procedures, often performed under spinal or combined spinal – epidural anesthesia. In these settings, spinal anesthesia itself is closely associated with rapid sympathetic blockade and post-spinal hypotension, which may complicate the interpretation of hemodynamic changes attributable to esketamine. By contrast, our study was conducted during emergency conversion from labor epidural analgesia to cesarean delivery, with anesthesia extended through a continuous epidural catheter. Compared with spinal anesthesia, epidural anesthesia generally produces a more gradual block and is less likely to cause abrupt hypotension. This feature may have reduced neuraxial anesthesia-related hemodynamic confounding and allowed a clearer assessment of whether low-dose esketamine meaningfully affects maternal blood pressure in a time-urgent bridging setting[31–32].Also,given the exceptionally unique and rare nature of this clinical scenario, in methodology, we designed to matched the statistical approach to the structure of each outcome. For repeated continuous variables, we used linear mixed-effects models with subject-specific random intercepts to account for within-patient correlation and between-patient heterogeneity. Baseline values were also included and centered, since the initial physiological state could continue to influence subsequent measurements and centering improved parameter interpretation, underscoring the need to account for individual hemodynamic status when estimating the independent effect of esketamine. For categorical repeated outcomes, we used generalized estimating equations to obtain population-averaged estimates, with generalized linear mixed models as sensitivity analyses. The consistency in effect direction and statistical interpretation across models further supported the robustness of the findings.

To our knowledge, this is one of the few clinical studies examining esketamine in the unique setting of Category I cesarean section. Although this special clinical scenario enhances the clinical relevance and novelty of our findings, it also introduces inherent limitations, most notably the restricted sample size. The modest sample size and the imbalance in group numbers may have reduced the power to detect small between-group differences or group-by-time interactions; therefore, some negative findings should be interpreted with caution.Second, measurements were obtained from baseline to 20 min after administration at 5-min intervals. Brief early hemodynamic peaks or transient adverse events related to esketamine may therefore have been missed.Third, to protect maternal safety, we excluded parturients with hypertension, cardiovascular disease, or the need for vasopressors or chronotropic agents. Patients requiring conversion to other anesthetic techniques were also not included. These criteria made the study population relatively stable, but they also limited the generalizability of the findings to higher-risk or hemodynamically unstable parturients and may have introduced selection bias. Obstetric patients are physiologically distinct from other surgical populations, supine hypotensive syndrome, rapid volume shifts, and dynamic perioperative changes may all influence maternal hemodynamics [6]. By excluding patients who required hemodynamic support, the study may have selectively retained more stable individuals.Fourth, maternal safety assessment in this study focused mainly on vital signs, pain, and sedation.

Neuropsychiatric symptoms such as dizziness, somnolence, or perceptual disturbances have been reported after perioperative ketamine or esketamine, including in emergency cesarean settings [12,20]. Because these events were not systematically quantified, future studies should include structured maternal adverse-event monitoring.Finally, although no immediate neonatal deterioration was observed in our study, early Apgar scores may be influenced by the underlying obstetric condition itself, such as fetal distress or placental abruption. Future studies should include more sensitive neonatal endpoints, including umbilical arterial blood gas analysis and NICU admission [11–12,24]. Larger multicenter studies involving higher-risk parturients, longer observation periods, different dosing regimens, and further evaluation of local anesthetic compatibility are warranted.In addition, participant recruitment began in September 2023, whereas the trial was retrospectively registered in April 2026 because the investigators became aware of the public trial registration requirement only after recruitment had begun. Although ethics approval was obtained before enrollment and institutional oversight continued throughout the study, these measures do not substitute for prospective registration. This delayed registration represents a limitation, and future clinical trials will be registered before participant enrollment.

## Conclusions

In emergency conversion from labor analgesia to Category I cesarean section, a highly time-sensitive setting, low-dose intravenous esketamine (0.15 mg/kg) used as an adjunct did not show an evident adverse effect on hemodynamics or oxygen saturation within the predefined 20-min observation window. Although SBP tended to increase slightly, neither the between-group difference nor the group-by-time interaction reached statistical significance. HR and SpO_2_ showed mild decreases over time, but again without significant treatment effects or interactions, suggesting that the short-term circulatory and respiratory effects of esketamine were generally manageable. No unfavorable signal was observed in Apgar scores, supporting its preliminary acceptability with respect to early neonatal safety [11,12,20].Esketamine showed clear clinical value in improving intraoperative comfort. It significantly reduced the likelihood of intraoperative pain and increased the probability of achieving the target sedation level. These findings suggest that, when neuraxial blockade has not yet been fully established but fetal delivery must proceed urgently, low-dose esketamine may provide a practical bridging analgesic-sedative strategy. Its main benefit lies in reducing pain, anxiety, and intraoperative discomfort while improving maternal cooperation and overall perioperative comfort [3,8,11–12,30].The findings also highlight the importance of baseline physiological status. Baseline SBP, HR, and SpO_2_ strongly influenced subsequent trajectories, indicating that interindividual heterogeneity and baseline differences should be carefully considered when evaluating drug effects in emergency cesarean research. Larger multicenter studies, especially those including higher-risk parturients, are still needed to clarify the optimal use and long-term safety of low-dose esketamine in this special clinical setting.

## Supporting information

supplementary table 3(data dictionary)

## Data Availability

All data produced in the present study are available upon reasonable request to the authors
All data produced in the present work are contained in the manuscript
All data produced are available online at

---

S1 Table. Definitions, analysis metrics, and timing of trial outcomes

For the repeated continuous outcomes, the 0-min measurement was considered the baseline value for the corresponding variable, including baseline systolic blood pressure, baseline heart rate, and baseline SpO_2_.

S2 Table. Sensitivity analysis of SpO2 using a beta generalized linear mixed-effects model(GLMM)

Notes:

1. Data are presented as odds ratios (ORs) with 95% confidence intervals (CIs) at each corresponding time point.
2. A beta generalized linear mixed-effects model (GLMM) with a logit link was fitted as a sensitivity analysis for SpO2.
3. Contrasts represent ke vs. ns at each time point; intervals are back-transformed from the log odds ratio scale.

## Notes

### Competing Interest Statement

The authors have declared no competing interest.

### Clinical Trial

ChiCTR2600121948

### Author Declarations

The study was reviewed and approved by the Ethics Committee of Foshan Nanhai District Maternal and Child Health Hospital before participant enrollment (approval No. 2022-01)

## Reference

[1] Anim-Somuah M, Smyth RM, Cyna AM, Cuthbert A. Epidural versus non-epidural or no analgesia for pain management in labour. Cochrane Database Syst Rev. 2018;5(5):CD000331. doi:10.1002/14651858.CD000331.pub4.

[2] Bloom SL, Leveno KJ, Spong CY, et al. Decision-to-incision times and maternal and infant outcomes. Obstet Gynecol. 2006;108(1):6–11. doi:10.1097/01.AOG.0000224693.07785.14

[3] Grap SM, Patel GR, Huang J, Vaida SJ. Risk factors for labor epidural conversion failure requiring general anesthesia for cesarean delivery. J Anaesthesiol Clin Pharmacol. 2022;38(1):118–123. doi:10.4103/joacp.JOACP_192_20.

[4] Arakawa M, Aoyama Y, Ohe Y. Epidural bolus injection with alkalinized lidocaine improves blockade of the first sacral segment--a brief report. Can J Anaesth. 2002;49(6):566–570. doi:10.1007/BF03017382

[5] Mendelson CL. The aspiration of stomach contents into the lungs during obstetric anesthesia. Am J Obstet Gynecol. 1946;52:191–205. doi:10.1016/s0002-9378(16)39829-5

[6] Chooi C, Cox JJ, Lumb RS, et al. Techniques for preventing hypotension during spinal anaesthesia for caesarean section. Cochrane Database Syst Rev. 2017;8(8):CD002251. Published 2017 Aug 4. doi:10.1002/14651858.CD002251.pub3

[7] Mion G, Villevieille T. Ketamine pharmacology: an update (pharmacodynamics and molecular aspects, recent findings). CNS Neurosci Ther. 2013;19(6):370–380. doi:10.1111/cns.12099.

[8] Gorlin AW, Rosenfeld DM, Ramakrishna H. Intravenous sub-anesthetic ketamine for perioperative analgesia. J Anaesthesiol Clin Pharmacol. 2016;32(2):160–167. doi:10.4103/0970-9185.182085.

[9] Zanos P, Moaddel R, Morris PJ, et al. Ketamine and ketamine metabolite pharmacology: insights into therapeutic mechanisms. Pharmacol Rev. 2018;70(3):621–660. doi:10.1124/pr.117.015198.

[10] Salahudeen MS, Wright CM, Peterson GM. Esketamine: new hope for the treatment of treatment-resistant depression? A narrative review. Ther Adv Drug Saf. 2020;11:2042098620937899. doi:10.1177/2042098620937899.

[11] Shen J, Song C, Lu X, et al. The effect of low-dose esketamine on pain and post-partum depression after cesarean section: a randomized controlled trial. Front Psychiatry. 2023;13:1038379. doi:10.3389/fpsyt.2022.1038379.

[12] Heesen M, Böhmer J, Klöhr S, et al. Intravenous ketamine during spinal and general anaesthesia for caesarean section: systematic review and meta-analysis. Acta Anaesthesiol Scand. 2015;59(4):414–426. doi:10.1111/aas.12468.

[13] Schulz KF, Altman DG, Moher D; CONSORT Group. CONSORT 2010 statement: updated guidelines for reporting parallel group randomized trials. BMJ. 2010;340:c332. doi:10.1136/bmj.c332.

[14] Whitehead AL, Julious SA, Cooper CL, Campbell MJ. Estimating the sample size for a pilot randomised trial to minimise the overall trial sample size for the external pilot and main trial for a continuous outcome variable. Stat Methods Med Res. 2016;25(3):1057–1073. doi:10.1177/0962280215588241

[15] Lucas DN, Yentis SM, Kinsella SM, et al. Urgency of caesarean section: a new classification. J R Soc Med. 2000;93(7):346–350. doi:10.1177/014107680009300703.

[16] Apgar V. A Proposal for a New Method of Evaluation of the Newborn Infant. Originally published in July 1953, volume 32, pages 250-259. Anesth Analg. 2015;120(5):1056–1059. doi:10.1213/ANE.0b013e31829bdc5c

[17] Ramsay MA, Savege TM, Simpson BR, Goodwin R. Controlled sedation with alphaxalone-alphadolone. Br Med J. 1974;2(5920):656–659.

[18] Hunger M, Döring A, Holle R. Longitudinal beta regression models for analyzing health-related quality of life scores over time. BMC Med Res Methodol. 2012;12:144. Published 2012 Sep 17. doi:10.1186/1471-2288-12-144

[19] Kinsella SM, Carvalho B, Dyer RA, et al. International consensus statement on the management of hypotension with vasopressors during caesarean section under spinal anaesthesia. Anaesthesia. 2018;73(1):71–92. doi:10.1111/anae.14080

[20] Huang XM, Qiu HX. Effect of intravenous esketamine in emergency cesarean deliveries: a retrospective analysis of maternal and neonatal outcomes. J Matern Fetal Neonatal Med. 2024;37(1):2413855. doi:10.1080/14767058.2024.2413855

[21] Kamp J, van Velzen M, Aarts L, Niesters M, Dahan A, Olofsen E. Stereoselective ketamine effect on cardiac output: a population pharmacokinetic/pharmacodynamic modelling study in healthy volunteers. Br J Anaesth. 2021;127(1):23–31. doi:10.1016/j.bja.2021.02.034

[22] Suleiman Z, Ik K, Bo B. Evaluation of the cardiovascular stimulation effects after induction of anaesthesia with ketamine. J West Afr Coll Surg. 2012;2(1):38–52.

[23] Zhang Y, Ye F, Zhang T, et al. Structural basis of ketamine action on human NMDA receptors. Nature. 2021;596(7871):301–305. doi:10.1038/s41586-021-03769-9

[24] Adhikari P, Subedi A, Sah BP, Pokharel K. Analgesic effects of intravenous ketamine after spinal anaesthesia for non-elective caesarean delivery: a randomised controlled trial. BMJ Open. 2021;11(6):e044168. Published 2021 Jun 30. doi:10.1136/bmjopen-2020-044168

[25] Regan KJ, O’Sullivan G. The extension of epidural blockade for emergency Caesarean section: a survey of current UK practice. Anaesthesia. 2008;63(2):136–142. doi:10.1111/j.1365-2044.2007.05319.x

[26] Popham P, Buettner A, Mendola M. Anaesthesia for emergency caesarean section, 2000-2004, at the Royal Women’s Hospital, Melbourne. Anaesth Intensive Care. 2007;35(1):74–79. doi:10.1177/0310057X0703500110

[27] Hillyard SG, Bate TE, Corcoran TB, Paech MJ, O’Sullivan G. Extending epidural analgesia for emergency Caesarean section: a meta-analysis. Br J Anaesth. 2011;107(5):668–678. doi:10.1093/bja/aer300

[28] Lam DT, Ngan Kee WD, Khaw KS. Extension of epidural blockade in labour for emergency Caesarean section using 2% lidocaine with epinephrine and fentanyl, with or without alkalinisation. Anaesthesia. 2001;56(8):790–794. doi:10.1046/j.1365-2044.2001.02058-4.x

[29] Zhu A, Pei L, Liu W, Cheng W, Zhang Y, Huang Y. Neurologic Complication Due to Crystallization After Drug Interaction Between Alkalized Lidocaine and Ropivacaine: A Case Report and in vitro Study. Front Med (Lausanne). 2022;9:919911. Published 2022 May 26. doi:10.3389/fmed.2022.919911

[30] Grap SM, Patel GR, Huang J, Vaida SJ. Risk factors for labor epidural conversion failure requiring general anesthesia for cesarean delivery. J Anaesthesiol Clin Pharmacol. 2022;38(1):118–123. doi:10.4103/joacp.JOACP_192_20

[31] Yang JR, Li YY, Ran TJ, et al. Esketamine Combined with Dexmedetomidine to reduce Visceral Pain During elective Cesarean Section Under Combined Spinal-Epidural Anesthesia: A double-Blind Randomized Controlled Study. Drug Des Devel Ther. 2024;18:2381–2392. Published 2024 Jun 18. doi:10.2147/DDDT.S460924

[32] Ng K, Parsons J, Cyna AM, Middleton P. Spinal versus epidural anaesthesia for caesarean section. Cochrane Database Syst Rev. 2004;2004(2):CD003765. doi:10.1002/14651858.CD003765.pub2

