## supplementary table 1 for "Clinical Effects of Low-Dose Esketamine During Emergency Conversion From Labor Epidural Analgesia to Category I Cesarean Section: A Randomized Controlled Trial of Maternal Hemodynamics, Analgesia, Sedation, and Neonatal Outcomes"

S1 Table

Definitions, analysis metrics, and timing of trial outcomes

| **Outcome** | **Variable** | **Analysis metric** | **Aggregation / summary** | **Time point** |
| --- | --- | --- | --- | --- |
| Primary outcome | Systolic blood pressure | Longitudinal change over time | β coefficient / adjusted mean difference with 95% CI | 0, 5, 10, 15, 20 min |
| Secondary outcome | Heart rate | Longitudinal change over time | β coefficient / adjusted mean difference with 95% CI | 0, 5, 10, 15, 20 min |
| Secondary outcome | SpO₂ | Longitudinal change over time | β coefficient / adjusted mean difference with 95% CI | 0, 5, 10, 15, 20 min |
| Secondary outcome | Intraoperative pain | Repeated binary outcome | OR with 95% CI | 5, 10, 15, 20 min |
| Secondary outcome | Surgeon satisfaction | Repeated binary outcome | OR with 95% CI | 5, 10, 15, 20 min |
| Secondary outcome | Ramsay target sedation | Single binary outcome | OR with 95% CI | 5 min |
| Secondary outcome | Apgar score | Descriptive neonatal score | Median, IQR, and counts | 1, 5, 10 min after birth |

**note：**

1. For the repeated continuous outcomes, the 0-min measurement was considered the baseline value for the corresponding variable, including baseline systolic blood pressure, baseline heart rate, and baseline SpO_2_ .
