## supplementary table 2 for "Clinical Effects of Low-Dose Esketamine During Emergency Conversion From Labor Epidural Analgesia to Category I Cesarean Section: A Randomized Controlled Trial of Maternal Hemodynamics, Analgesia, Sedation, and Neonatal Outcomes"

S1 Table

Sensitivity analysis of SpO2 using a beta generalized linear mixed-effects model (GLMM)

| Outcome / Predictor | OR | 95% CI | P-value |
| --- | --- | --- | --- |
| Time 5 min: ke vs. ns | 1.353 | [0.863, 2.12] | 0.187 |
| Time 10 min: ke vs. ns | 0.910 | [0.594, 1.40] | 0.666 |
| Time 15 min: ke vs. ns | 0.796 | [0.525, 1.21] | 0.282 |
| Time 20 min: ke vs. ns | 0.986 | [0.657, 1.48] | 0.945 |

Notes:

1. Data are presented as odds ratios (ORs) with 95% confidence intervals (CIs).
2. A beta generalized linear mixed-effects model (GLMM) with a logit link was fitted as a sensitivity analysis for SpO2.
3. Contrasts represent ke vs. ns at each time point; intervals are back-transformed from the log odds ratio scale.
