## supplementary table 3(data dictionary) for "Clinical Effects of Low-Dose Esketamine During Emergency Conversion From Labor Epidural Analgesia to Category I Cesarean Section: A Randomized Controlled Trial of Maternal Hemodynamics, Analgesia, Sedation, and Neonatal Outcomes"

### Data dicrionary

| Variable name | Description | Type / format | Values / coding | Unit / notes |
|---|---|---|---|---|
| id | Participant identifier | Integer | Unique de-identified participant ID | Each participant has one unique ID |
| drug | Treatment group | Categorical | ns = normal saline group; ke = esketamine group | Group assignment |
| time | Measurement time point | Numeric | 0, 5, 10, 15, 20 | Minutes after study drug administration or start of observation window |
| bp | Systolic blood pressure | Continuous numeric | Recorded value | mmHg |
| rate | Heart rate | Continuous numeric | Recorded value | beats/min |
| hurt | Intraoperative pain | Binary categorical | 0 = no pain; 1 = pain | Recorded at repeated intraoperative time points |
| ramsay | Ramsay sedation score | Binary categorical | 0 = not target sedation; 1 = target sedation | Ramsay sedation scale score; in analysis, scores 2–4 were coded as target sedation |
| an_satisf | Surgeon satisfaction with muscle relaxation | Binary categorical | 0 = not satisfied; 1 = satisfied | Repeated intraoperative assessment |
| bbapgar | Neonatal Apgar score | Ordinal / discrete numeric | 0–10 | Apgar score assessed at 1, 5, and 10 min after birth |
| wei | Maternal body weight | Continuous numeric | Recorded value | kg |

| | | | | |
|---|---|---|---|---|
| age | Maternal age | Continuous numeric | Recorded value | years |
| spo2 | Peripheral oxygen saturation | Continuous numeric | Recorded value | % |
